# Automated Echocardiographic Detection of Congenital Heart Disease Using Artificial Intelligence

**DOI:** 10.64898/2026.01.24.26344771

**Authors:** Platon Lukyanenko, Sunil Ghelani, Yuting Yang, Bohan Jiang, Timothy Miller, David Harrild, Nao Sasaki, Francesca Sperotto, Danielle Sganga, John Triedman, Andrew J. Powell, Tal Geva, William G. La Cava, Joshua Mayourian

## Abstract

**Background:** Delayed or missed diagnosis of congenital heart disease (CHD) contributes to excess pediatric mortality worldwide. Echocardiography (echo) is central to diagnosing and triaging CHD, yet expert interpretation remains a scarce and maldistributed global resource. Artificial intelligence (AI) offers the potential to democratize diagnostics and extend expert-level interpretation beyond large academic centers, but its application in CHD remains underexplored.

**Methods:** We developed EchoFocus-CHD, an AI-enabled model for automated detection of 12 critical and 8 non-critical CHD lesions, individually and as composites. The composite critical CHD outcome was the primary endpoint. The model expands on a multi-task, view-agnostic architecture (PanEcho) with a transformer encoder to improve focus on relevant echo views. The model was trained (80%) and tested (20%) on the first echo per patient from Boston Children’s Hospital (BCH), with external validation on US and international studies from patients referred to BCH.

**Results:** The internal and external cohorts included 3.4 million videos from 54,727 echos (median age at echo 7.1 [IQR, 0.2-15.0] years; 5.8% critical CHD; 23.6% non-critical CHD) and 167,484 videos from 3,356 echos (median age at echo 2.5 [IQR, 0.3-9.4] years; 29.4% critical CHD; 45.6% non-critical CHD), respectively. EchoFocus-CHD showed excellent internal ability to detect the composite critical CHD outcome (AUROC 0.94, LR+ 7.50, LR- 0.14) and individual critical lesions (AUROC 0.83-1.00), as well as composite non-critical CHD (AUROC 0.90, LR+ 5.00, LR- 0.23) and individual non-critical lesions (AUROC 0.70-0.96). Performance declined during external validation to detect critical CHD (AUROC 0.77), coinciding with greater expert disagreement on external cases (κ=0.72 versus 0.82 for internal cases). Explainability analyses demonstrated that the model prioritized the same clinically relevant views (parasternal long-axis, parasternal short-axis, and subxiphoid long-axis) across internal and external cohorts, while UMAP analysis revealed a domain shift between cohorts. Retraining on all available US patients attenuated domain shift, improving international critical CHD detection (AUROC 0.87) and calibration.

**Conclusions:** EchoFocus-CHD shows promise for automated CHD detection and highlights the need to address domain shift for real-world deployment. By identifying high-risk CHD lesions, this approach could support triage, prioritize expert review, and optimize resource allocation, advancing more equitable global cardiovascular care.

## INTRODUCTION

Congenital heart disease (CHD) affects approximately 1 in 100 live births, impacting over 12 million individuals worldwide.^1,2^ Nearly 25% of CHD cases are critical, often requiring urgent intervention in the neonatal period to prevent cardiovascular collapse and death.^3^ Unfortunately, CHD is frequently diagnosed late in both low-resource^4^ and high-resource^5^ countries, reflecting a persistent diagnostic gap. This challenge is particularly severe in low- and middle-income countries (LMICs) where the burden of disease is greatest^6^ and access to diagnostics and congenital care are limited,^6,7^ highlighting the global imperative for timely and effective CHD detection and triage.

Echocardiography (echo) is the cornerstone of pediatric cardiology and CHD diagnosis, providing non-invasive, real-time assessment of cardiac anatomy and function without radiation. Pediatric echo interpretation is technically challenging: it requires the interpretation of complex, heterogeneous lesions in small hearts and is often complicated by motion artifacts and variable image quality. These challenges are compounded by a global shortage of pediatric cardiologists and specialized imaging experts,^6–8^ creating a critical bottleneck for timely and accurate diagnoses.

Artificial intelligence (AI) has shown promise to address diagnostic gaps in adult echo. For example, AI-echo models can reliably automate measurements,^9–12^ assess heart muscle and valve function,^13^ or even provide a comprehensive echo evaluation.^14^ In contrast, transthoracic AI-echo for pediatric cardiology remains nascent, with prior work largely limited to view classification,^15^ isolated measurement tasks,^16,17^ or detection of specific findings (e.g., patent ductus arteriosus)^18^ rather than comprehensive structural screening.^19^

To address this technological gap, we developed EchoFocus-CHD, a multi-task, view-agnostic AI-echo model designed to automatically detect a broad spectrum of critical and non-critical CHD lesions. To evaluate performance under real-world conditions and assess generalizability, we externally validated the model using echos from 58 countries across 6 continents, with the goal of enabling scalable CHD triage and prioritization in resource-limited settings.

## METHODS

This study is reported in accordance with the TRIPOD+AI 2024 guidelines.^20^

### Patient Population and Patient Assignment

Patient data and echos were obtained from Boston Children’s Hospital (BCH) between July 2015 and July 2025. Only transthoracic echos with ≥10 DICOM files were included in this study; fetal echos and echos performed in the operating room were excluded. Echos that did not pass quality control criteria (see “Data Retrieval, Pre-Processing, and Quality Control” below) were also excluded. Given our objective to identify previously unknown or unverified CHD, only the first echo per patient was included. These criteria defined the main study cohort.

The main cohort was subsequently partitioned into internal studies (performed at BCH, Brigham and Women’s Hospital nursery/NICU, Beth Israel nursery/NICU, or affiliated BCH satellite clinics) and external studies (outside referral echos read by BCH expert cardiac imagers for diagnostic assistance or second opinions). The external cohort was further subdivided into US and international patients. International patients were defined as having non-US home addresses. Within the internal cohort, patients were randomly assigned in an 80:20 ratio to development and testing cohorts.

### Definition of Outcomes

Diagnostic labels for each echo were derived from the Fyler coding system—a detailed, decades-old, well-established anatomic classification system used at BCH and specifically designed for CHD.^21^ For every echo, expert interpreting cardiac imagers (with sub-specialty training in non-invasive pediatric cardiac imaging) assign Fyler codes that capture both major and minor structural cardiac lesions with high anatomic granularity.

Outcomes of interest included critical and non-critical CHD lesions, predicted individually and as composites (Table S1). The composite critical CHD outcome was the primary endpoint. Outcome labels were not mutually exclusive (i.e., a patient can have tetralogy of Fallot and an atrial septal defect).

CHD lesions were considered as critical if surgical or catheter-based intervention is typically required within the first year of life. The 12 individual critical CHD lesions predicted were double outlet right ventricle, D-loop transposition of the great arteries, Ebstein anomaly, hypoplastic left heart syndrome, tricuspid atresia, truncus arteriosus, any functional single ventricle lesions (broadly defined as “single ventricle”, “single left ventricle”, or “single right ventricle”), tetralogy of Fallot, atrioventricular canal defect, coarctation of the aorta, pulmonary atresia, and totally anomalous pulmonary venous connection. The composite critical CHD outcome indicates the presence of any of these individual lesions, in addition to anomalous left coronary artery from the pulmonary artery, aortopulmonary window, double-outlet left ventricle, interrupted aortic arch, critical aortic stenosis, and critical pulmonary stenosis (Table S1). These additional lesions were not predicted individually due to insufficient positive samples.

A CHD lesion was considered non-critical if it is typically managed conservatively or with intervention delayed beyond infancy. The 8 non-critical CHD lesions predicted were atrial septal defect, anomalies of coronary artery origins, bicuspid aortic valve, left superior vena cava, partially anomalous pulmonary venous connection, ductus arteriosus, right aortic arch, and ventricular septal defect. The composite non-critical CHD outcome indicates the presence of any of these individual lesions, in addition to the following less common non-critical lesions that were not predicted individually: cor triatriatum, double aortic arch, [I,D,D] transposition of the great arteries, left pulmonary artery sling, [S,L,L] transposition of the great arteries, and vascular ring.

### Data Retrieval, Pre-Processing, and Quality Control

All echocardiographic studies were retrieved from the institutional picture archiving and communication system (PACS). All echos underwent pre-processing analogous to that described in the PanEcho framework.^9^

Pixel data from two-dimensional echo videos were first extracted from DICOM files. All videos then underwent comprehensive deidentification. Specifically, each frame was binarized using a fixed threshold, and all pixels outside the convex hull of the largest detected contour were masked. Videos were subsequently cropped to the central image content in a temporally consistent manner, downsampled to a resolution of 256 x 256 pixels using bicubic interpolation, and further deidentified by masking peripheral regions containing protected health information.^9^

### EchoFocus-CHD Model Architecture

The EchoFocus-CHD architecture takes a set of echo videos from a single study as input and produces multiple task-specific predictions of CHD classifications. The architecture extends PanEcho^9^ by adapting the final layers of the network with additional transformer layers to allow attention^22^ to operate over video clip embeddings (Figure 1B). Analogous to how a human expert interprets an echo, the attention mechanism enables the model to selectively weight diagnostically informative videos, enhancing the representation of relevant structural and functional features for CHD classification.

**Figure 1:**
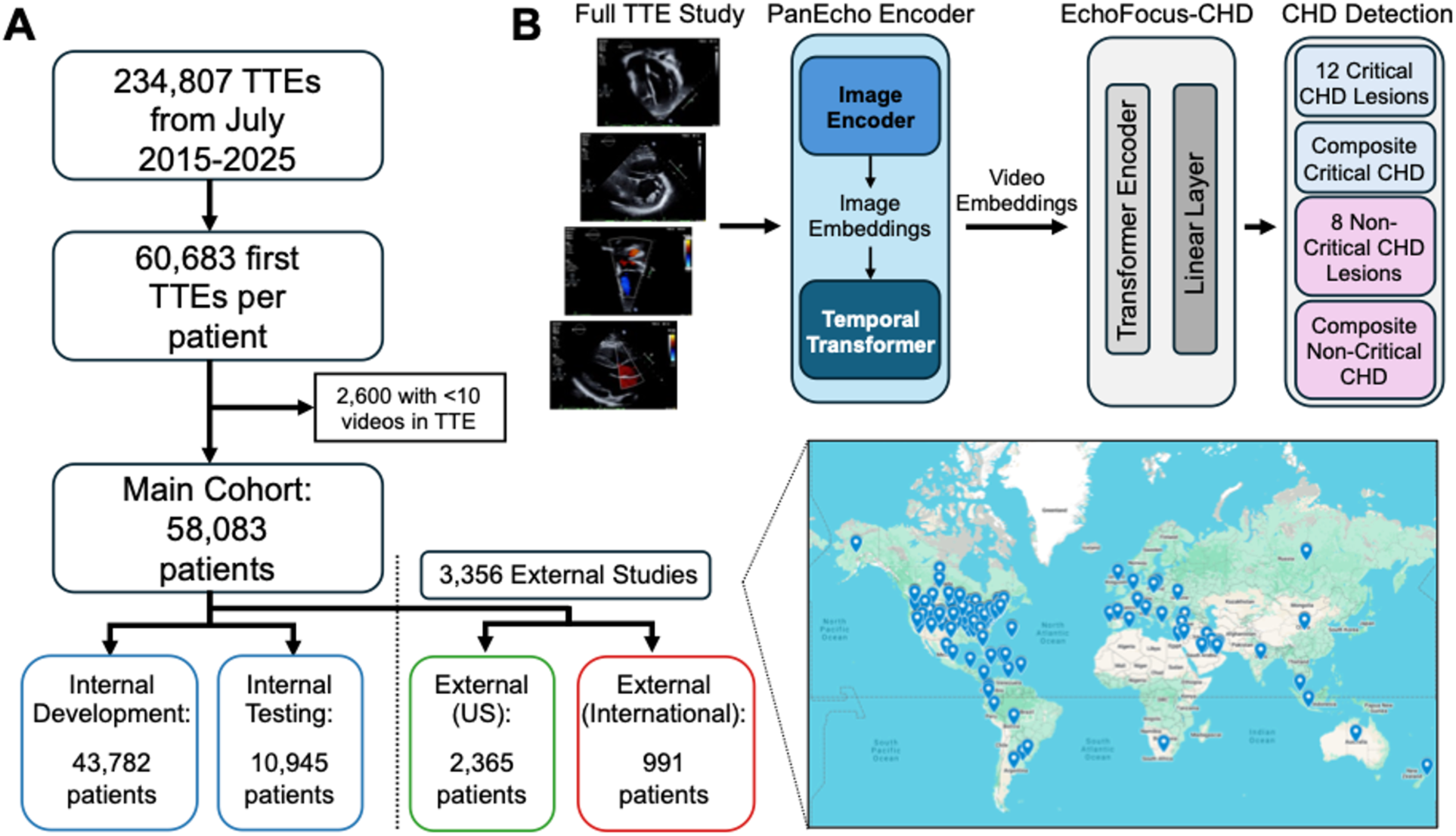
Schematic of Study Design and Model Architecture. (**A**) Schematic of training and testing design. STROBE diagram showing initial patient selection and filtering at each data processing stage (with primary outcome rates shown). Pins of origin countries for outside patients inset. (**B**) Schematic of EchoFocus-CHD architecture and classification targets. Abbreviations: transthoracic echo (TTE).

Echo videos are first separated into 16 random sets of 16 sequential frames (called clips); each frame (image) is individually processed with a 2D convolutional neural network (ConvNeXt-T,^23^ pretrained on ImageNet) to produce image embeddings. These image embeddings are stacked sequentially and fed into a temporal transformer, consisting of 4 layers with 8 attention heads. This process mimics the use of transformers for interpreting natural language sentences; in this setting, the image embeddings are like word “tokens”, and the clips are treated as “sentences”. To capture the temporal information of the frames, a standard positional encoding is added to the image tokens. For each clip in the echo study, the output of the temporal transformer is aggregated using mean pooling to produce a clip-level embedding, represented as a 768-dimensional vector.

EchoFocus-CHD then departs from the PanEcho architecture^9^ by introducing a study-level transformer encoder that operates across all (number of videos x 16) clip-level embeddings to generate a single study-level embedding. This transformer encoder leverages self-attention to learn additional dependencies between videos in the study before moving to task prediction. The resulting study-level embedding is then passed through fully connected layers to generate task-specific outputs of CHD classification labels.

### Model Training

The internal BCH cohort designated for model development was randomly partitioned into training (80%) and validation (20%) sets. The model was trained using the training set, with the validation set used exclusively for model selection. During training, pretrained PanEcho model weights were frozen and used to generate video-level embeddings, allowing optimization to focus on learning the parameters of the study-level transformer encoder and the fully connected, task-specific output layers.

Training was performed using the AdamW optimizer^24^ with a weight decay of 0.01 and a scheduled learning rate that decreased upon plateaus in validation loss. Training was terminated after 10 consecutive epochs without improvement in validation loss.

Several strategies were employed to improve training robustness. Consistent with the PanEcho approach,^9^ we utilized several image augmentation techniques (cropping, rotation, and flipping), to improve robustness to imaging noise. On layers following PanEcho, dropout^25^ was applied during training at a rate of 0.2 with an additional clip-level dropout at 0.5 to enhance robustness to missing video clips.

For hyperparameter tuning, we varied the depth of the study-level transformer encoder (1, 5, 10, and 20 layers), the learning rate (0.0001-0.01), and the effective batch size (32-128). The final model was selected by minimizing loss across tasks on the held-out validation set.

### Model Performance Evaluation and Statistical Analyses

Model discrimination was assessed using the area under the receiver operating characteristic curve (AUROC). Additional clinically relevant performance metrics included sensitivity, specificity, positive and negative predictive values, positive and negative likelihood ratios (LRs), and lift. These metrics were computed using decision thresholds that maximize the Youden index, derived from the validation set. Confidence intervals for performance metrics were estimated using 1,000 bootstrap samples.

Descriptive data are presented as frequencies and percentages for categorical variables and median and interquartile range (IQR) for continuous variables.

### Model Calibration Analysis

Model calibration was assessed via calibration plots and scaled Brier scores. Scaled Brier scores measure the mean squared difference between predicted probabilities and observed outcomes, scaled relative to the score of a non-informative model predicting the cohort’s outcome prevalence. This scaling accounts for differences in outcome prevalence across cohorts and provides an interpretable metric ranging from 0 (no improvement over baseline) to 1 (perfect prediction).

### Sensitivity and Subgroup Analyses

We evaluated the model’s robustness for detecting exclusively unrepaired CHD through a sensitivity analysis that excluded echos from patients with prior cardiac interventions (i.e., catheterization or surgery), as determined by Fyler codes. To assess sensitivity to outcome labeling, we compared model performance when using structured Fyler code labels versus labels automatically extracted from echo report text by an internal instance of GPT-4o-mini (OpenAI, San Francisco, CA).

Subgroup analyses were performed on the test cohorts stratified by age and number of echo videos per study. Age groupings were adapted from prior work^26^ and defined as age < 1 (infant), 1-3, 3-8, 8-12, 12-18 years, and age >18 years. Echo videos per study groupings were defined as <25, 26-50, 51-75, 76-100, and >100. Model discrimination within each age subgroup was assessed using AUROC.

### Model Adjudication

Four expert cardiac imagers characterized model errors through an adjudication process: for both internal and external studies, 2 experts each independently reviewed 25 random false positive and 25 random false negative infant echos. Adjudicators reviewed the full echo study and were blinded to patient name, echo report, model predictions, and to each other’s assessments. For each echo, adjudicators were asked to classify the study into one of 4 categories: 1) critical CHD; 2) non-critical CHD; 3) indeterminate (due to inadequate image quality); or 4) indeterminate (due to evolving physiology requiring follow-up, such as suspected coarctation of the aorta in the presence of a ductus arteriosus). Adjudication outcomes between internal and external cohorts were compared using the Fisher’s exact test.

For the purposes of evaluating agreement in a triage context, we calculated Cohen’s kappa (κ) when grouping indeterminate studies with non-critical studies to yield a binary critical versus non-critical/indeterminate classification. A Cohen’s κ value of 1 indicates perfect agreement, 0 indicates agreement equivalent to chance, and values less than 0 indicate agreement worse than chance.

### Model Explainability

To interpret model predictions, an integrated gradients-based explainability analysis was performed for one left-sided lesion (hypoplastic left heart syndrome) and one right-sided lesion (tetralogy of Fallot). For each lesion, we selected 25 internal and 25 external echo studies with positive cases and the smallest prediction errors. For each echo study, integrated gradients were applied to quantify the contribution of individual video clips to the model’s predicted output.

The 10 most highly weighted video clips per study were identified and subsequently reviewed by an expert cardiac imager, who recorded: 1) which unique echo views the model prioritized; 2) whether the 5 or 10 highest prioritized video clips were sufficient to detect the lesion of interest.

### Embedding Visualization for Domain Shift Assessment

To explore potential domain shift^27^ (i.e., differences in training versus deployment echo imaging conditions that can degrade performance) between internal and external echo studies, we applied unsupervised Uniform Manifold Approximation and Projection (UMAP) on high-dimensional embeddings produced by the EchoFocus-CHD study-level transformer encoder. We applied UMAP using 15 neighbors and the cosine distance metric. The resulting space was visualized and qualitatively compared between internal and external cohorts to assess overlap and separation that might indicate domain shift related to differences in acquisition setting, patient population, or imaging protocols.

### Data Availability and Software

The model and source code are available from https://echofocus.org for non-commercial, academic-only purposes to accelerate research on AI-echo in pediatric cardiology. Requests for BCH data and related materials will be internally reviewed to clarify if the request is subject to intellectual property or confidentiality constraints. Shareable data and materials will be released under a material transfer agreement for non-commercial research purposes. Use of BCH data was approved by its Institutional Review Board.

## RESULTS

### Patient Population Characteristics

From the 234,807 transthoracic echos at Boston Children’s Hospital meeting inclusion criteria, 60,683 were first time studies per patient. After excluding echos with <10 DICOM files per study (n=2,600), there were 58,083 studies remaining, forming the main cohort (Figure 1A). Of those, 54,727 were internal studies and 3,356 studies were sent from outside centers: 2,365 from patients across the US, and 991 from international patients. International patients resided in 58 countries spanning 6 continents: North America, South America, Europe, Asia, Africa, and Australia.

As shown in Table 1, there were numerous differences between the internal and external cohorts. There were 2.6 million, 0.8 million, and 0.2 million videos within the internal development, internal testing, and outside cohorts, respectively (Table 1). The internal studies had more videos per study (median 75) compared to outside studies (median 46). Internal studies were performed at an older age (median age at echo 7.1 [IQR, 0.2-15.0] years) compared to external studies (median age at echo 2.5 [IQR, 0.3-9.4] years). There was a substantially higher prevalence of CHD in the external cohort (29.4% critical CHD; 45.6% non-critical CHD) compared to the internal cohort (5.8% critical CHD; 23.6% non-critical CHD). For details of prevalence for individual lesions within each cohort, see Table 1.

**Table 1:** Baseline Characteristics of Internal and External Cohorts.

|  | Internal Cohort |  | Outside Studies |  |  |
| --- | --- | --- | --- | --- | --- |
|  | Development | Testing | External (US) | External (International) | External (Combined) |
| Patients | 43782 | 10945 | 2365 | 991 | 3356 |
| TTEs | 43782 | 10945 | 2365 | 991 | 3356 |
| Videos | 2617348 | 818487 | 119152 | 48332 | 167484 |
| Videos Per Study | 75 (63, 87) | 75 (63, 88) | 45 (31, 65) | 47 (30, 65) | 46 (31, 65) |
| Age at TTE | 7.11 (0.25,14.96) | 6.96 (0.20,14.99) | 2.17 (0.34,8.96) | 3.3 (0.33,10.40) | 2.46 (0.33,9.36) |
| Sex (Male) | 23291 (53.20%) | 5752 (52.55%) | 1295 (54.76%) | 529 (53.38%) | 1824 (54.35%) |
| **Composite Critical CHD** | 2525 (5.77%) | 628 (5.74%) | 810 (34.25%) | 177 (17.86%) | 987 (29.41%) |
| ALCAPA | 7 (0.02%) | 2 (0.02%) | 3 (0.13%) | 0 (0.00%) | 3 (0.09%) |
| AP window | 13 (0.03%) | 5 (0.05%) | 7 (0.30%) | 0 (0.00%) | 7 (0.21%) |
| DORV | 163 (0.37%) | 35 (0.32%) | 116 (4.90%) | 30 (3.03%) | 146 (4.35%) |
| D-loop TGA | 235 (0.54%) | 62 (0.57%) | 55 (2.33%) | 17 (1.72%) | 72 (2.15%) |
| Ebstein | 83 (0.19%) | 25 (0.23%) | 54 (2.28%) | 17 (1.72%) | 71 (2.12%) |
| HLHS | 194 (0.44%) | 42 (0.38%) | 70 (2.96%) | 24 (2.42%) | 94 (2.80%) |
| IAA | 33 (0.08%) | 16 (0.15%) | 10 (0.42%) | 2 (0.20%) | 12 (0.36%) |
| Tricuspid Atresia | 82 (0.19%) | 13 (0.12%) | 11 (0.47%) | 7 (0.71%) | 18 (0.54%) |
| Truncus Arteriosus | 48 (0.11%) | 13 (0.12%) | 17 (0.72%) | 4 (0.40%) | 21 (0.63%) |
| SV Disease | 330 (0.75%) | 70 (0.64%) | 113 (4.78%) | 41 (4.14%) | 154 (4.59%) |
| Tetralogy of Fallot | 515 (1.18%) | 104 (0.95%) | 86 (3.64%) | 16 (1.61%) | 102 (3.04%) |
| AVCD | 406 (0.93%) | 109 (1.00%) | 254 (10.74%) | 35 (3.53%) | 289 (8.61%) |
| CoA | 806 (1.84%) | 203 (1.85%) | 131 (5.54%) | 21 (2.12%) | 152 (4.53%) |
| TAPVC | 71 (0.16%) | 13 (0.12%) | 20 (0.85%) | 8 (0.81%) | 28 (0.83%) |
| Critical AS | 10 (0.02%) | 2 (0.02%) | 4 (0.17%) | 1 (0.10%) | 5 (0.15%) |
| Critical PS | 26 (0.06%) | 8 (0.07%) | 2 (0.08%) | 0 (0.00%) | 2 (0.06%) |
| Pulmonary Atresia | 244 (0.56%) | 55 (0.50%) | 85 (3.59%) | 26 (2.62%) | 111 (3.31%) |
| **Composite Non-Critical CHD** | 10277 (23.47%) | 2619 (23.93%) | 1232 (52.09%) | 298 (30.07%) | 1530 (45.59%) |
| ASD | 3037 (6.94%) | 771 (7.04%) | 441 (18.65%) | 95 (9.59%) | 536 (15.97%) |
| Anomalous Coronaries | 302 (0.69%) | 69 (0.63%) | 49 (2.07%) | 6 (0.61%) | 55 (1.64%) |
| BAV | 1212 (2.77%) | 287 (2.62%) | 212 (8.96%) | 35 (3.53%) | 247 (7.36%) |
| Cor Triatriatum | 23 (0.05%) | 4 (0.04%) | 5 (0.21%) | 1 (0.10%) | 6 (0.18%) |
| Double Aortic Arch | 82 (0.19%) | 17 (0.16%) | 10 (0.42%) | 0 (0.00%) | 10 (0.30%) |
| Left PA sling | 15 (0.03%) | 3 (0.03%) | 4 (0.17%) | 1 (0.10%) | 5 (0.15%) |
| LSVC | 574 (1.31%) | 141 (1.29%) | 85 (3.59%) | 13 (1.31%) | 98 (2.92%) |
| L-loop TGA | 61 (0.14%) | 19 (0.17%) | 68 (2.88%) | 27 (2.72%) | 95 (2.83%) |
| PAPVC | 266 (0.61%) | 63 (0.58%) | 40 (1.69%) | 10 (1.01%) | 50 (1.49%) |
| PDA | 4521 (10.33%) | 1238 (11.31%) | 198 (8.37%) | 94 (9.49%) | 292 (8.70%) |
| Right Aortic Arch | 478 (1.09%) | 103 (0.94%) | 67 (2.83%) | 9 (0.91%) | 76 (2.26%) |
| Vascular Ring | 125 (0.29%) | 31 (0.28%) | 14 (0.59%) | 0 (0.00%) | 14 (0.42%) |
| VSD | 3177 (7.26%) | 794 (7.25%) | 461 (19.49%) | 95 (9.59%) | 556 (16.57%) |

Data presented as frequency (percentage) and median (interquartile range).
**Abbreviations:** anomalous left coronary artery from the pulmonary artery (ALCAPA); aortopulmonary (AP); bicuspid aortic valve (BAV); double outlet right ventricle (DORV); transposition of the great arteries (TGA); hypoplastic left heart syndrome (HLHS); interrupted aortic arch (IAA); left superior vena cava (LSVC); partial anomalous pulmonary venous connection (PAPVC); patent ductus arteriosus (PDA); single ventricle (SV); atrioventricular canal defect (AVCD); ventricular septal defect (VSD); coarctation of the aorta (CoA); total anomalous pulmonary venous connection (TAPVC); aortic/pulmonary stenosis (AS/PS).

### EchoFocus-CHD Model Performance

Model performance metrics of EchoFocus-CHD for individual critical CHD lesions during internal and external testing are shown in Figure 2 and Tables S1-S4. During internal testing, performance was excellent for a majority of lesions: AUROC 0.97 for Ebstein anomaly; AUROC ≥0.99 for single ventricle lesions such as hypoplastic left heart syndrome, tricuspid atresia, and any single ventricle lesion; AUROC ≥0.97 for conotruncal lesions such as double outlet right ventricle, D-loop transposition of the great arteries, truncus arteriosus, and tetralogy of Fallot; AUROC 0.96 for atrioventricular canal defects and pulmonary atresia; AUROC 0.90 for coarctation of the aorta; and AUROC 0.83 for total anomalous pulmonary venous connection. In comparison, there was a reduction in performance for the overall external cohort across all individual critical CHD lesions, with AUROC ranging from 0.70-0.85 (Figure 2).

**Figure 2:**
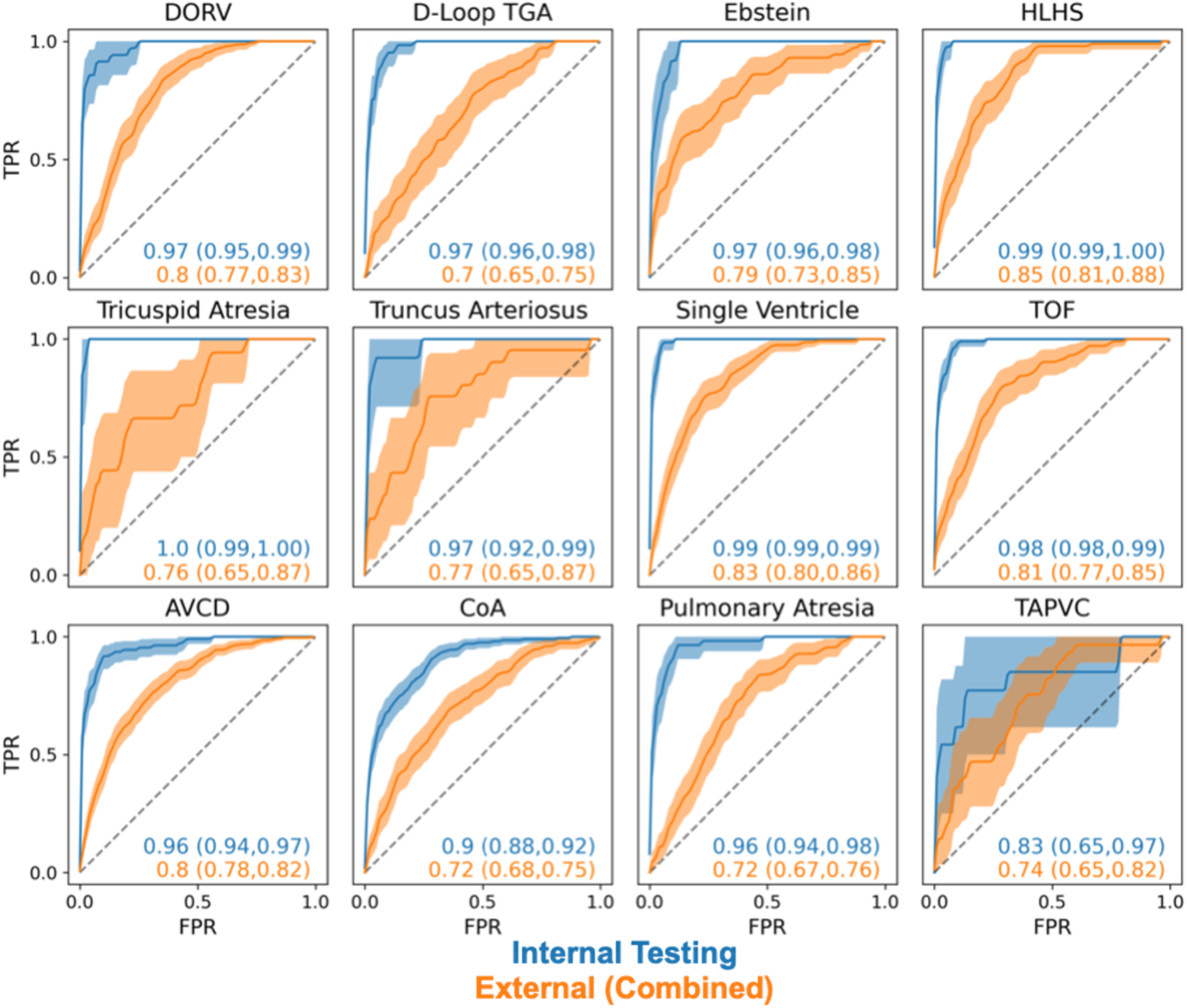
EchoFocus-CHD Performance to Predict Individual Critical CHD Lesions. Performance of EchoFocus-CHD to predict individual critical CHD lesions evaluated using the internal (blue) and overall external (orange) test cohorts using receiver operating curves. Dotted line represents chance. 95% confidence intervals are computed using bootstrapping. **Abbreviations:** true positive rate (TPR); false positive rate (FPR); double outlet right ventricle (DORV); transposition of the great arteries (TGA); hypoplastic left heart syndrome (HLHS); tetralogy of Fallot (TOF); atrioventricular canal defect (AVCD); coarctation of the aorta (CoA); total anomalous pulmonary venous connection (TAPVC).

For individual non-critical CHD lesions, internal performance ranged from AUROC 0.70 (anomalous coronaries) to 0.96 (ductus arteriosus). For atrial and ventricular septal wall defects, AUROC was 0.87 and 0.91, respectively (Table S2). Externally, performance also declined for non-critical CHD lesions. For example, external AUROC decreased to 0.80 for patent ductus arteriosus, 0.74 for atrial septal defect, and 0.72 for ventricular septal defect. Tables S2-S5 list performance metrics for individual non-critical CHD lesions.

When assessing the composite critical CHD outcome (Figure 3), internal performance was excellent in both the overall internal cohort (AUROC 0.94) and the infant subgroup (AUROC 0.93). In contrast, performance was lower in the external cohort (AUROC 0.77 for all external studies, 0.74 for US external studies, and 0.82 for international external studies), which further declined for the infant cohort (AUROC 0.71 for all external studies, 0.68 for US external studies, and 0.73 for international external studies). Calibration analysis (Figure S1) showed a moderate scaled Brier score of 0.405 for the internal cohort, whereas the external cohorts exhibited poor calibration, with scaled Brier scores of 0.045 for the overall external cohort, 0.005 for the external US cohort, and 0.067 for the external international cohort.

**Figure 3:**
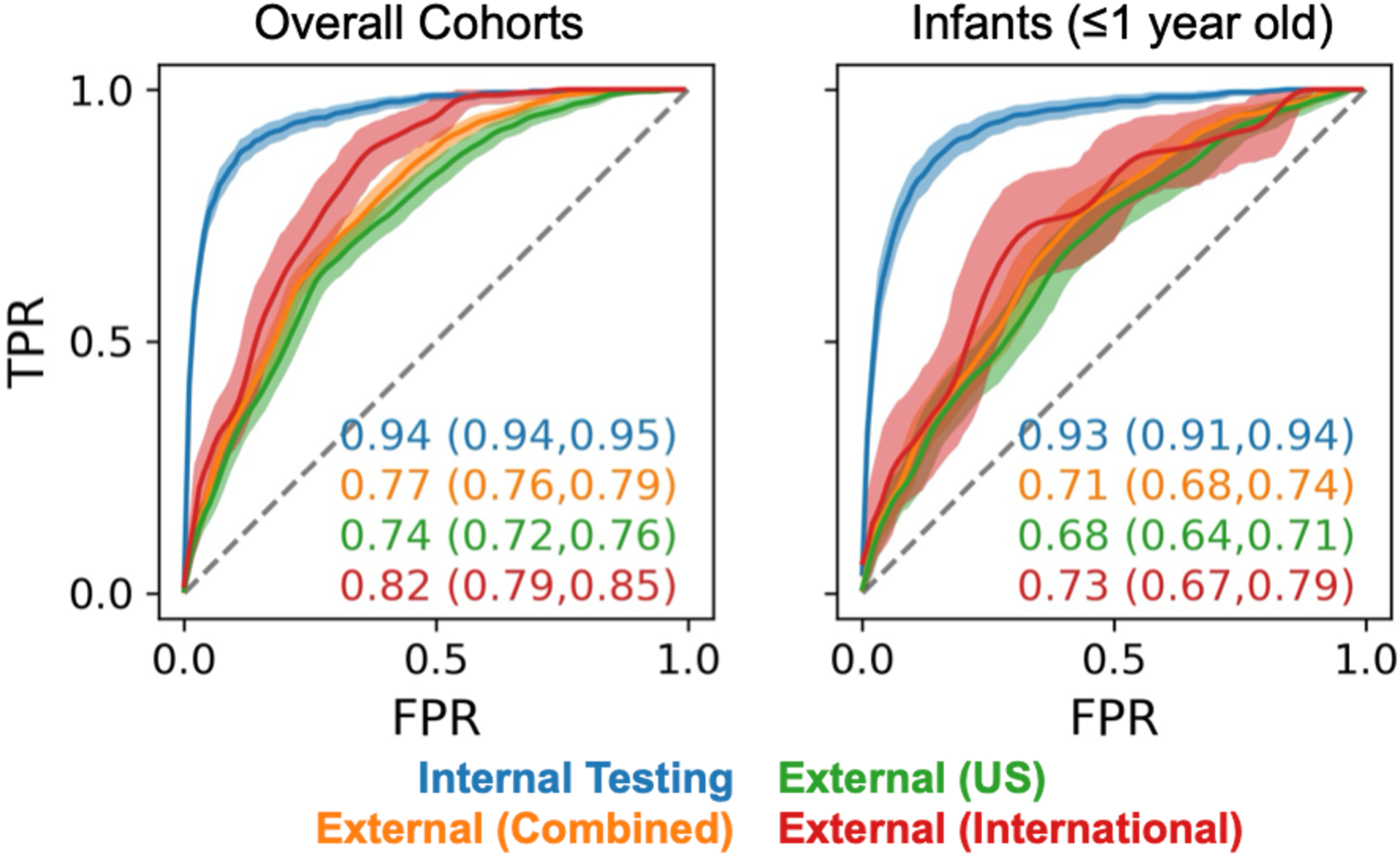
EchoFocus-CHD Performance to Predict the Composite Critical CHD Outcome. Performance of EchoFocus-CHD to predict the composite critical CHD outcome evaluated in the overall cohort (left) and infant subgroup (right) using the internal (blue), overall external (orange), external US (green), and external international (red) cohorts using receiver operating curves. Dotted line represents chance. 95% confidence intervals are shown using bootstrapping. **Abbreviations:** true positive rate (TPR); false positive rate (FPR); United States (US).

### Subgroup and Sensitivity Analyses

During sensitivity analysis, model performance to detect the composite critical CHD outcome was unchanged when excluding echos with prior cardiac interventions (internal AUROC 0.94 [95% CI, 0.93-0.95]; external AUROC 0.74 [95% CI, 0.72-0.76]). In addition, using labels generated by a large language model from echo report free text did not alter model performance (Table S6).

Subgroup analyses by study size demonstrated lower performance for critical CHD detection in studies with fewer than 25 videos (Table S7), whereas no consistent performance trends were observed across age subgroups (Table S8).

### Expert Adjudication

Expert adjudication was performed on 50 internal and 50 external discrepant test cases for both false negatives and false positives. For false negatives, adjudicators determined that 42% of internal cases were in fact negative, compared with 12% of external cases (Figure 4A). For false positives, adjudicators determined that 10% of internal cases were indeed positive, compared with 20% of external cases. The distributions of adjudication outcomes differed significantly between internal and external cohorts for both false negatives (p<0.001) and false positives (p=0.01). Inter-rater agreement was high internally (Cohen’s κ=0.82) with a drop to κ=0.72 externally, suggesting greater diagnostic ambiguity in the external cohort.

**Figure 4.**
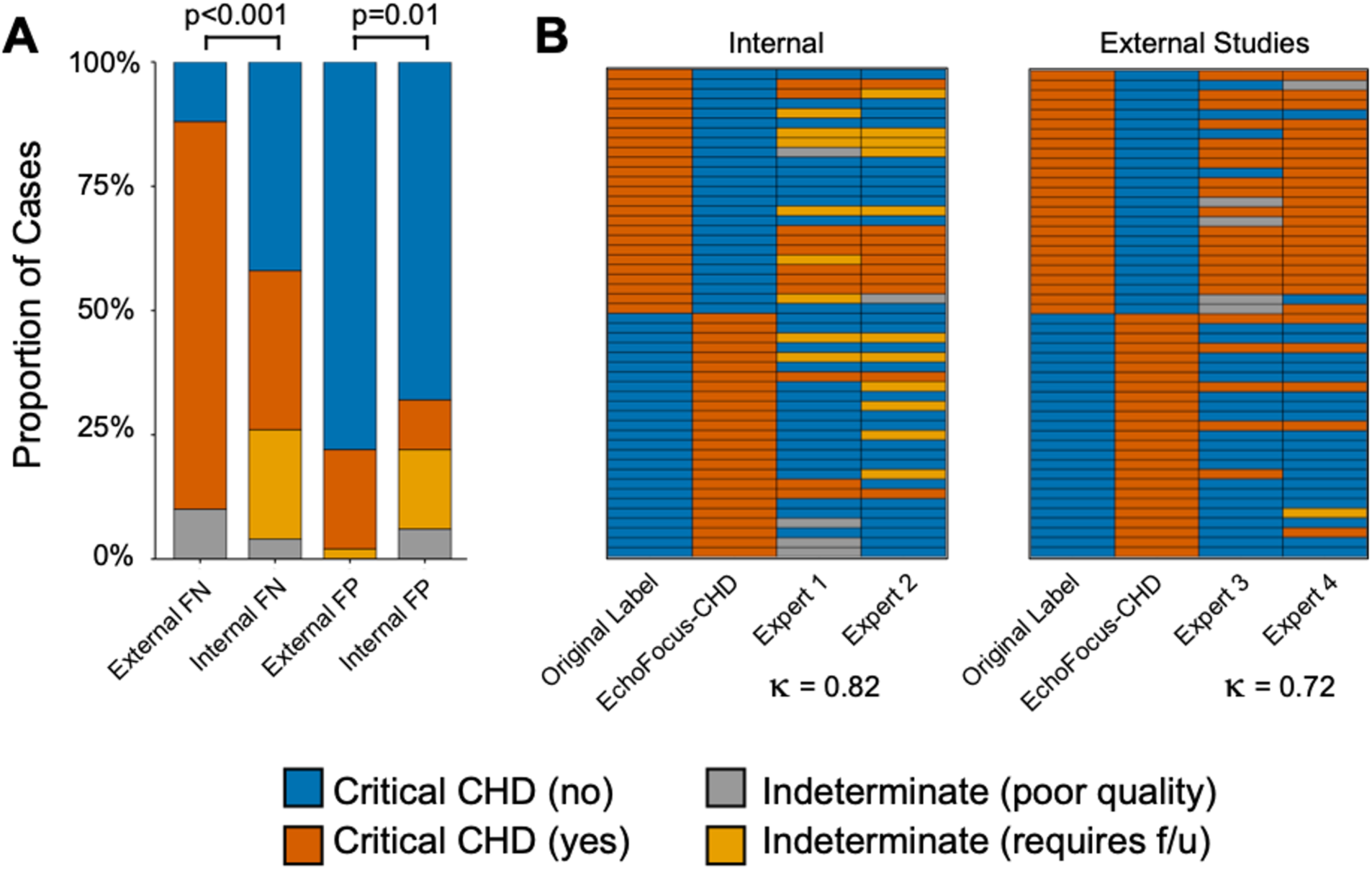
Expert Adjudication of Discrepant Cases. Expert adjudication was performed on 50 internal and 50 external discrepant test cases. (**A**) Stacked bar plot showing the proportion of cases classified as no critical CHD (blue), critical CHD (amber), indeterminate due to poor image quality (gray), and indeterminate due to evolving physiology requiring follow-up (yellow). P-value obtained via Fisher’s exact test. (**B**) Heatmap displaying study-level classifications assigned by each expert adjudicator; inter-rater agreement, assessed using Cohen’s κ, is inset. **Abbreviations:** congenital heart disease (CHD); false negative (FN); false positive (FP).

### Model Explainability Analysis

Across both internal and external test cohorts, model explainability consistently prioritized the same views to detect hypoplastic left heart syndrome and tetralogy of Fallot: parasternal long-axis, parasternal short-axis, and subxiphoid long-axis views (Figure 5A). In the majority of studies (range of 76-100%), the top 5 and top 10 attention-weighted clips were sufficient for an expert cardiac imager to determine the presence or absence of critical CHD (Figure 5B). There was no significant difference between internal and external cohorts in the ability to identify critical CHD from these clips.

**Figure 5.**
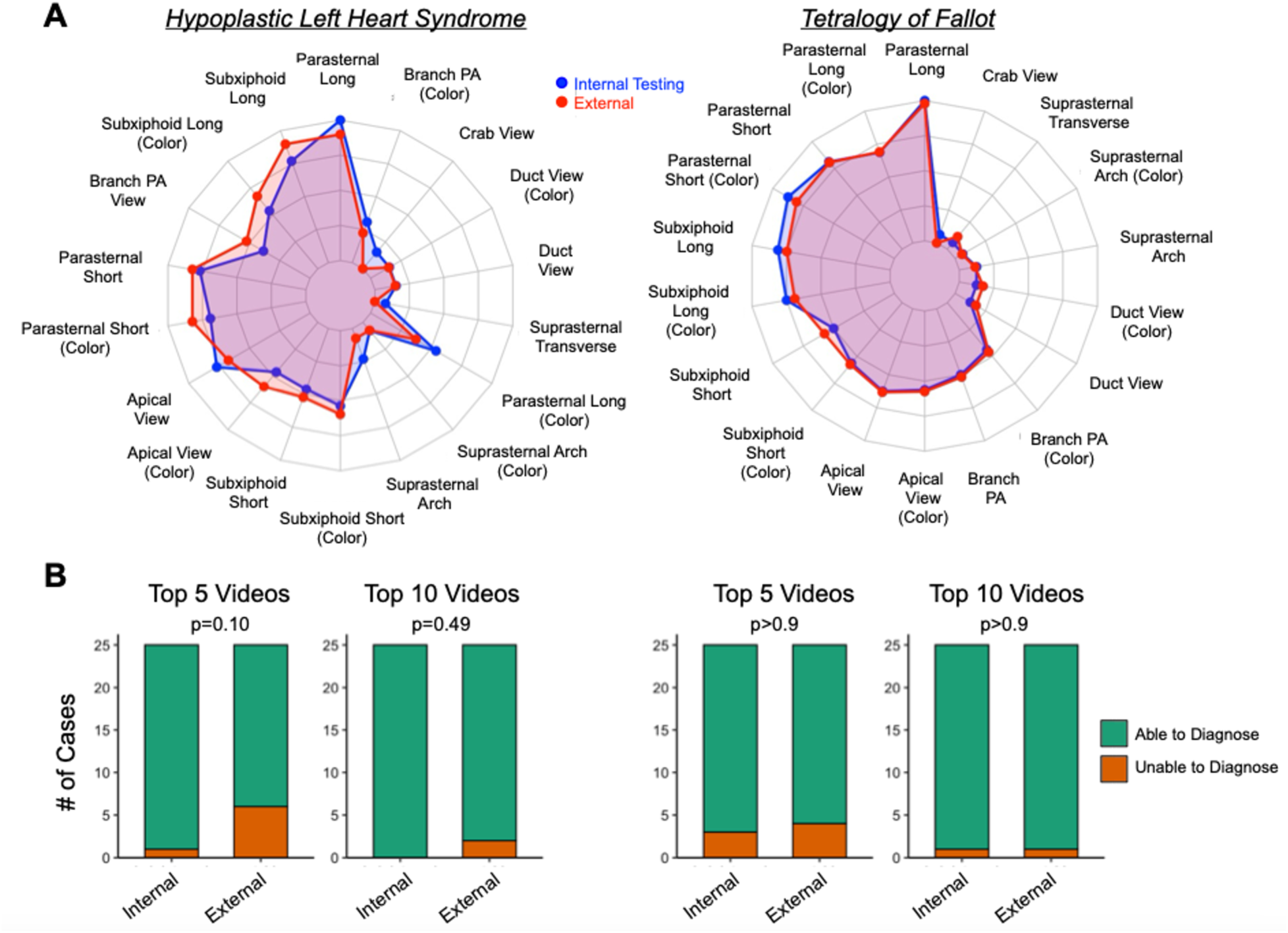
Expert Review of EchoFocus-CHD Model Attention for Diagnosing Critical CHD. (**A**) Radar plots of selected views in top 10 clips for hypoplastic left heart syndrome (left) and tetralogy of Fallot (right) for internal (blue) and external (red) studies. (**B**) Diagnostic accuracy of top EchoFocus-CHD selected clips. Stacked bar plots show the proportion of studies in which an expert imager could identify hypoplastic left heart syndrome (left) and tetralogy of Fallot (right) from the top 5 and top 10 clips selected by the model. P-values obtained via Fisher’s exact test. **Abbreviations:** pulmonary artery (PA).

### Exploring and Addressing Domain Shift

To explore whether domain shift contributed to lower external performance, we visualized study-level embeddings using UMAP. As shown in Figure S2A, internal studies formed several dense clusters, which only partially overlapped with the external clusters. Notably, some external studies occupied regions of the embedding space that were sparsely populated by internal studies, suggesting the presence of domain shift. This is particularly evident in the bottom right quadrant, where there was a high density of external critical CHD (Figure S2B).

To address domain shift, we retrained EchoFocus-CHD using an expanded model development cohort that incorporated all US external studies in addition to the original BCH training set. The BCH test cohort and the external international cohort were excluded from training. As shown in Figure 6, internal model performance for the composite critical CHD outcome remained excellent and largely unchanged across the overall cohort, infants, and individual critical CHD lesions (Table S9). Internal calibration was also unchanged (Figure S3). In the external international cohort, performance for the composite critical CHD outcome improved with AUROCs of 0.87 for the overall cohort and 0.84 for infants (Figure 6). External calibration also improved to 0.151 (Figure S3). For 10 of 12 individual critical CHD lesions, AUROC increased by a median of 0.08.

**Figure 6:**
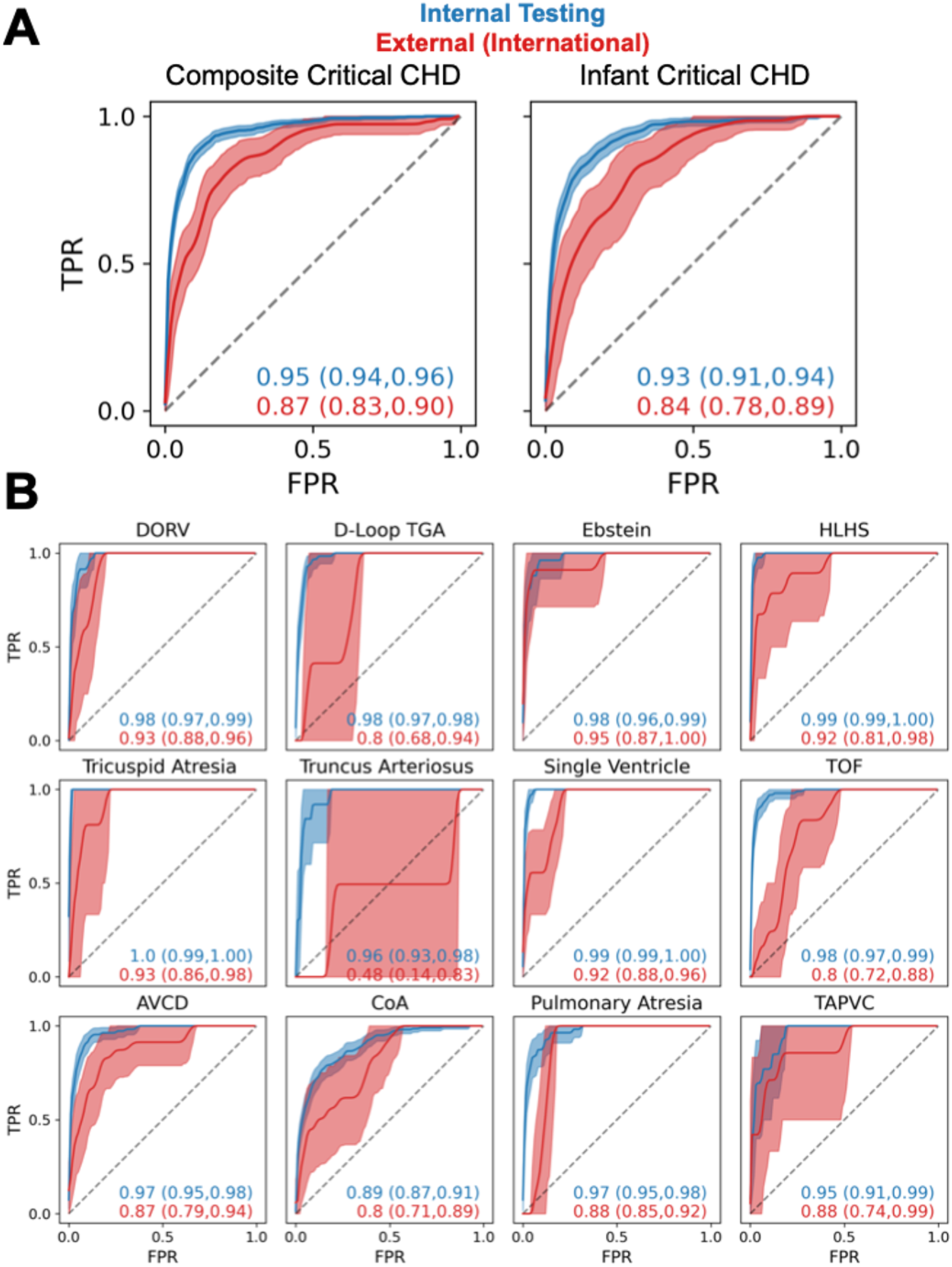
Retraining EchoFocus-CHD on Broader Dataset Improves Performance to Predict Critical CHD. Performance of retrained EchoFocus-CHD model to predict (**A**) the composite critical CHD outcome and (**B**) individual critical CHD outcomes on the internal (blue) and external international (red) cohorts using receiver operating curves. Dotted line represents chance. 95% confidence intervals are computed using bootstrapping. **Abbreviations:** true positive rate (TPR); false positive rate (FPR); double outlet right ventricle (DORV); transposition of the great arteries (TGA); hypoplastic left heart syndrome (HLHS); tetralogy of Fallot (TOF); atrioventricular canal defect (AVCD); coarctation of the aorta (CoA); total anomalous pulmonary venous connection (TAPVC).

Across the retrained internal versus external cohorts, sensitivity was similar (86-88%), while specificity was lower in the external international cohort (72% versus 89%). The negative LR was comparable between cohorts (0.13-0.19), whereas the positive LR was higher in the internal cohort (7.8 vs. 3.2). Across internal and external cohorts, positive predictive values were 33% [95% CI, 32-34%] and 35% [95% CI, 30-41%], respectively; negative predictive values were 99% [95% CI, 99-99%] and 97% [95% CI, 96-98%], respectively. Full external international performance metrics are provided in Table S10.

## DISCUSSION

In this study, we developed a view-agnostic, multi-task AI-echo model for automated detection of a broad spectrum of CHD lesions. The model introduces a novel study-level transformer encoder as an extension of the PanEcho^9^ framework, enabling integration of information across multiple video clips in a manner analogous to how cardiologists synthesize findings across views, highlighting both the architectural innovation and clinical plausibility of this approach. Using the largest pediatric echo dataset reported to date, we demonstrate excellent internal discrimination for both composite and individual CHD outcomes. We further evaluate the model’s external generalizability across a large, geographically diverse external referral cohort, identifying performance degradation partly attributable to domain shift and demonstrating that discrimination and calibration can be improved through retraining with more heterogeneous data. Expert adjudication revealed lower inter-rater agreement externally among pediatric cardiologists, suggesting that external cases missed by the model may represent diagnostically challenging studies rather than unequivocal errors. Altogether, EchoFocus-CHD illustrates the potential of AI-echo to function as a clinical decision-support tool, prioritizing and triaging studies in resource-limited settings to optimize timely access to scarce pediatric cardiology and congenital surgery expertise, rather than serving as a replacement for clinician interpretation.

### Global Disparities in Pediatric Cardiology Care

There is an underrecognized global burden of pediatric heart disease,^28^ with CHD constituting a leading cause of childhood non-communicable mortality worldwide.^1^ It is estimated that more than 90% of children born with CHD reside in LMICs, which together account for 94% of global CHD-related mortality.^28,29^ Even in more developed nations, CHD related mortality is higher in rural and more resource-constrained regions.^30^ Reducing these inequities is therefore central to achieving the United Nations’ Sustainable Development Goals targeting reductions in neonatal and under-five mortality by 2030.^31^

Despite this urgency, many pediatric cardiac care systems remain fragile, driven in large part by critical shortages of clinicians with specialized expertise in the diagnosis and management of pediatric heart disease.^28^ For example, most countries in sub-Saharan Africa and many in Asia lack structured training programs in pediatric cardiology and congenital cardiac surgery^31^ and facilities capable of performing infant or neonatal cardiac surgery.^32^ Existing models of pediatric heart care in high-income countries are unfeasible for LMICs, requiring alternative and context-appropriate strategies to facilitate timely referral to specialized centers. Similar challenges and proposed solutions have been described in rural and underserved regions of high-income countries such as the US.

Within this framework, EchoFocus-CHD was developed as an initial step toward enabling scalable, technology-assisted CHD screening and prioritization, with the goal of extending limited pediatric cardiology expertise to settings where it is most constrained.

### Clinical Implications of EchoFocus-CHD

EchoFocus-CHD is intended to function as a triage and decision-support tool in resource-constrained settings, where access to pediatric cardiology expertise is limited and timely prioritization of high-risk patients is critical. In this context, the model’s operating characteristics support clinically meaningful risk stratification. Internally, EchoFocus-CHD demonstrated high sensitivity and specificity (both ∼90%), corresponding to strong positive and negative LRs (7.8 and 0.13, respectively; Table S9). Externally, sensitivity remained high (0.86) with moderate specificity (0.72), yielding a preserved negative LR of 0.13 and a positive LR of 3.2 (Table S10). These findings indicate that the model is particularly effective for ruling out critical CHD (with negative predictive values of 97-99% across cohorts), a key requirement for triage applications in which false negatives carry substantial clinical risk. In addition, approximately one-third of cases flagged as positive by EchoFocus-CHD were confirmed to be critical CHD (i.e., positive predictive value of 33-35% across cohorts).

Notably, the performance metrics likely underestimate true clinical accuracy, as adjudication identified a subset of cases initially labeled as incorrect that were either correct, evolving physiology (e.g., suspected coarctation of the aorta in the setting of a patent ductus arteriosus), diagnostically ambiguous, or challenging even for expert readers. Importantly, EchoFocus-CHD demonstrated good internal calibration, with improved calibration in the retrained external international cohort. In low-resource environments where downstream resources such as specialist consultation, transport, or advanced imaging are limited, well-calibrated risk estimates may allow for rational prioritization rather than reliance on binary classification alone. Beyond binary triage, EchoFocus-CHD provides lesion-specific subtype predictions, which may further inform urgency, anticipated clinical course, and referral pathways.

### Importance of Real-World Deployment

A central objective of this study was to evaluate model performance in a large, diverse cohort that was geographically and demographically distinct from the training population, reflecting conditions expected during real-world deployment. Model performance declined in the external cohort (Figure 3), independent of outcome labeling approach (i.e., Fyler versus large language model), number of videos per study, or differences in patient age. While top selected views were consistent across cohorts (Figure 5A) and clinically relevant (Figure 5B), there were discernable differences in the model representations between internal and external datasets (Figure S2). Domain shift—the phenomenon where a model’s performance degrades when applied to data that differ from its training set—is anticipated in pediatric echo, a modality characterized by substantial variability in vendor-specific image processing, operator-dependent acquisition techniques, image quality, and institution-specific protocols. These factors introduce meaningful heterogeneity that must be carefully considered as AI-based echo tools move toward clinical deployment.

To help disentangle two plausible sources of domain shift in this study (underlying patient population versus echo acquisition), we incorporated external US referral echos into the training set. The observed improvement in external international performance following this retraining step suggests that a component of the generalization gap is attributable to differences in image acquisition/processing rather than solely to population-level differences. This finding highlights the importance of dataset heterogeneity, particularly with respect to imaging practices, for improving model robustness.

### Limitations and Future Directions

Several limitations merit consideration. First, despite retraining on a more heterogeneous US cohort, performance on the external international cohort remains below the threshold for safe clinical deployment. This highlights the ongoing need to improve model generalizability, which could be addressed through strategies such as: 1) exploring alternative/hybrid architectures (e.g., EchoPrime)^14^ or learning approaches (e.g., adversarial learning^33^); 2) developing a pediatric and CHD-specific foundation model to generate a more robust embedding space; 3) leveraging multi-institutional or federated learning approaches to incorporate data from both large and small centers;^34^ and 4) multi-modal approaches,^35^ such as integrated AI-enabled ECG.^36–38^ Second, although model performance was comparable when using either Fyler-coded labels or large language model-extracted labels from echo reports, both approaches are imperfect. Fyler codes are highly granular but may be affected by human documentation limitations, while large language model-extracted labels are prone to misinterpretation of report text. Consequently, labeling errors may persist. Third, although our external validation set was geographically diverse, certain regions of particular clinical interest (most notably sub-Saharan Africa) were not represented, potentially limiting the generalizability of findings to areas with the greatest unmet need. Fourth, our models rely on transthoracic echos acquired by trained sonographers; translation to low-resource or point-of-care settings will require validation on portable ultrasound studies, which may have lower image quality and greater operator variability. Fifth, while the model encompasses a broad spectrum of lesions, it does not provide predictions for all pediatric heart conditions (e.g., Kawasaki disease, rheumatic heart disease, cardiomyopathy). Finally, while integrated gradients-based explainability was performed, further work is needed to evaluate how these visualizations impact clinician trust and decision-making in practice.

Future directions should include continued model refinement for low-resource settings, prospective multi-site evaluation in diverse healthcare environments, and formal assessment of clinical utility and workflow integration.

## Conclusions

EchoFocus-CHD demonstrates that large-scale, multi-task AI models show promise to detect a wide range of CHD lesions from routine echo. At the same time, our findings highlight the critical importance of external validation, calibration assessment, and domain shift mitigation for real-world implementation. By identifying both strengths and limitations, this work provides a foundation for future prospective studies and iterative deployment strategies to advance equitable, scalable CHD care worldwide.

## ACKNOWLEDGMENTS

The authors would like to acknowledge Boston Children’s Hospital’s High-Performance Computing Resources Clusters Enkefalos 3 (E3) made available for conducting the research reported in this publication.

## SUPPLEMENTAL MATERIAL

Tables S1-S10

Figures S1-S3

## Nonstandard Abbreviations and Acronyms

AI: Artificial Intelligence
AUROC: Area under the Receiver Operating Curve
BCH: Boston Children’s Hospital
CHD: Congenital Heart Disease
Echo: Echocardiography
LMIC: Low- and Middle-Income Countries
LR: Likelihood Ratio
UMAP: Uniform Manifold Approximation and Projection

