## Supplementary Materials for "Automated Echocardiographic Detection of Congenital Heart Disease Using Artificial Intelligence"

**Running Title:** AI-Echo to Detect Congenital Heart Disease

Platon Lukyanenko*,[a] Sunil Ghelani*,[b] Yuting Yang,[a] Bohan Jiang,[a] Timothy Miller,[a] David Harrild,[b] Nao Sasaki,[b] Francesca Sperotto,[b] Danielle Sganga, [b] John Triedman,[b] Andrew J. Powell,[b] Tal Geva,[b] William G. La Cava**,[a], Joshua Mayourian**,[b]

[a] Computational Health Informatics Program, Boston Children's Hospital, Department of Pediatrics, Harvard Medical School, Boston, MA, USA
[b] Department of Cardiology, Boston Children's Hospital, Department of Pediatrics, Harvard Medical School, Boston, MA, USA

* Co-First Authors
** Co-Senior Authors

**Address for correspondence:**
Joshua Mayourian
Department of Cardiology, Boston Children's Hospital
300 Longwood Avenue
Boston, MA 02115


**Table S1: Critical and Non-Critical Lesion Definitions**

| Individually Predicted Critical CHD Lesions | Composite Critical CHD Outcome | Individually Predicted Non-Critical CHD Lesions | Composite Non-Critical CHD Outcome |
|---|---|---|---|
| DORV | ALCAPA | ASD | ASD |
| D-loop TGA | AP window | Anomalous Coronaries | Anomalous Coronaries |
| Ebstein | DORV | BAV | BAV |
| HLHS | D-loop TGA | LSVC | Cor Triatriatum |
| Tricuspid Atresia | Ebstein | PAPVC | Double Aortic Arch |
| Truncus Arteriosus | HLHS | PDA | Left PA sling |
| SV Disease | IAA | Right Aortic Arch | LSVC |
| Tetralogy of Fallot | Tricuspid Atresia | VSD | L-loop TGA |
| AVCD | Truncus Arteriosus | | PAPVC |
| CoA | SV Disease | | PDA |
| TAPVC | Tetralogy of Fallot | | Right Aortic Arch |
| Pulmonary Atresia | AVCD | | Vascular Ring |
| | CoA | | VSD |
| | TAPVC | | |
| | Critical AS | | |
| | Critical PS | | |
| | Pulmonary Atresia | | |
| | DOLV | | |

### Table S2: Internal Model Performance for Each Outcome

| | AUROC | Sensitivity | Specificity | LR + | LR - | Lift |
|---|---|---|---|---|---|---|
| ASD | 0.87 (0.86,0.89) | 0.83 (0.81,0.86) | 0.78 (0.77,0.79) | 3.75 (3.57,3.95) | 0.22 (0.19,0.25) | 5.74 (5.25,6.30) |
| Anomalous Coronaries | 0.7 (0.63,0.76) | 0.6 (0.48,0.72) | 0.66 (0.65,0.67) | 1.77 (1.43,2.14) | 0.61 (0.42,0.78) | 4.24 (2.22,7.37) |
| BAV | 0.85 (0.82,0.87) | 0.74 (0.69,0.79) | 0.8 (0.79,0.80) | 3.65 (3.37,3.93) | 0.32 (0.26,0.39) | 12.63 (10.45,14.82) |
| DORV | 0.97 (0.95,0.99) | 0.86 (0.74,0.96) | 0.95 (0.95,0.96) | 18.54 (15.57,21.56) | 0.15 (0.04,0.28) | 61.76 (35.86,101.23) |
| D-loop TGA | 0.97 (0.96,0.98) | 0.95 (0.89,1.00) | 0.89 (0.88,0.89) | 8.51 (7.90,9.10) | 0.05 (0.00,0.12) | 60.96 (40.54,84.99) |
| Ebstein | 0.97 (0.96,0.98) | 1.0 (1.00,1.00) | 0.86 (0.85,0.87) | 7.11 (6.80,7.46) | 0.0 (0.00,0.00) | 70.08 (31.39,131.62) |
| HLHS | 0.99 (0.99,1.00) | 0.95 (0.88,1.00) | 0.95 (0.95,0.96) | 20.78 (18.48,23.02) | 0.05 (0.00,0.13) | 137.58 (95.31,199.43) |
| LSVC | 0.9 (0.87,0.92) | 0.83 (0.77,0.88) | 0.82 (0.81,0.82) | 4.53 (4.15,4.88) | 0.21 (0.14,0.29) | 15.19 (10.97,20.26) |
| PAPVC | 0.85 (0.80,0.90) | 0.71 (0.60,0.82) | 0.81 (0.80,0.81) | 3.66 (3.05,4.25) | 0.36 (0.22,0.50) | 22.25 (12.28,36.99) |
| PDA | 0.96 (0.96,0.97) | 0.91 (0.90,0.93) | 0.88 (0.87,0.89) | 7.59 (7.18,8.01) | 0.1 (0.08,0.12) | 7.08 (6.75,7.45) |
| Right Aortic Arch | 0.87 (0.83,0.91) | 0.78 (0.70,0.86) | 0.83 (0.82,0.84) | 4.62 (4.09,5.11) | 0.27 (0.17,0.37) | 14.78 (9.39,21.54) |
| Tricuspid Atresia | 1.0 (0.99,1.00) | 0.85 (0.62,1.00) | 0.99 (0.99,1.00) | 140.47 (96.85,188.21) | 0.15 (0.00,0.38) | 398.91 (183.11,771.64) |
| Truncus arteriosus | 0.97 (0.92,0.99) | 0.92 (0.71,1.00) | 0.93 (0.92,0.93) | 13.0 (10.06,14.81) | 0.09 (0.00,0.31) | 64.41 (25.26,135.48) |
| SV Disease | 0.99 (0.99,0.99) | 0.97 (0.93,1.00) | 0.95 (0.95,0.96) | 20.15 (18.35,22.19) | 0.03 (0.00,0.08) | 93.03 (70.70,121.87) |
| Tetralogy of Fallot | 0.98 (0.98,0.99) | 0.96 (0.92,0.99) | 0.93 (0.92,0.93) | 12.84 (11.90,13.89) | 0.04 (0.01,0.08) | 48.71 (38.30,61.99) |
| AVCD | 0.96 (0.94,0.97) | 0.93 (0.88,0.97) | 0.87 (0.86,0.88) | 7.19 (6.69,7.68) | 0.08 (0.03,0.14) | 41.56 (32.18,52.41) |
| VSD | 0.91 (0.90,0.92) | 0.86 (0.84,0.88) | 0.8 (0.79,0.80) | 4.24 (4.04,4.43) | 0.17 (0.15,0.20) | 7.53 (6.96,8.11) |
| CoA | 0.9 (0.88,0.92) | 0.71 (0.65,0.77) | 0.89 (0.88,0.89) | 6.21 (5.57,6.80) | 0.33 (0.26,0.40) | 14.99 (12.00,18.53) |
| Pulmonary Atresia | 0.96 (0.94,0.98) | 0.89 (0.80,0.97) | 0.91 (0.91,0.92) | 10.18 (9.10,11.33) | 0.12 (0.04,0.22) | 62.22 (39.44,93.40) |
| TAPVC | 0.83 (0.65,0.97) | 0.62 (0.33,0.88) | 0.91 (0.91,0.92) | 7.07 (3.79,10.10) | 0.42 (0.14,0.73) | 78.95 (15.93,203.19) |
| Composite Non-Critical CHD | 0.9 (0.89,0.91) | 0.81 (0.80,0.83) | 0.84 (0.83,0.85) | 5.0 (4.74,5.28) | 0.23 (0.21,0.24) | 3.35 (3.25,3.46) |
| Composite Critical CHD | 0.94 (0.94,0.95) | 0.88 (0.85,0.90) | 0.88 (0.88,0.89) | 7.5 (7.05,7.96) | 0.14 (0.11,0.16) | 10.95 (10.11,11.83) |
| Infant Composite Critical CHD | 0.93 (0.91,0.94) | 0.93 (0.91,0.96) | 0.74 (0.73,0.76) | 3.64 (3.41,3.90) | 0.09 (0.06,0.12) | 6.13 (5.59,6.73) |

**Table S3: External (Combined) Model Performance for Each Outcome**

| | AUROC | Sensitivity | Specificity | LR + | LR - | Lift |
|---|---|---|---|---|---|---|
| ASD | 0.74 (0.72,0.77) | 0.74 (0.70,0.78) | 0.62 (0.60,0.64) | 1.94 (1.81,2.07) | 0.42 (0.36,0.48) | 2.26 (2.03,2.51) |
| Anomalous Coronaries | 0.74 (0.67,0.81) | 0.64 (0.51,0.78) | 0.7 (0.68,0.71) | 2.12 (1.68,2.56) | 0.51 (0.32,0.70) | 5.68 (2.97,10.26) |
| BAV | 0.75 (0.72,0.78) | 0.71 (0.65,0.76) | 0.65 (0.63,0.66) | 2.01 (1.83,2.18) | 0.45 (0.37,0.54) | 4.08 (3.34,4.91) |
| DORV | 0.8 (0.77,0.83) | 0.57 (0.49,0.65) | 0.82 (0.81,0.83) | 3.19 (2.67,3.71) | 0.53 (0.42,0.63) | 3.32 (2.56,4.40) |
| D-loop TGA | 0.7 (0.65,0.75) | 0.64 (0.52,0.74) | 0.63 (0.61,0.65) | 1.72 (1.38,2.02) | 0.58 (0.41,0.77) | 2.84 (1.79,4.50) |
| Ebstein | 0.79 (0.73,0.85) | 0.69 (0.59,0.79) | 0.72 (0.71,0.74) | 2.48 (2.08,2.88) | 0.43 (0.29,0.58) | 6.99 (4.18,10.76) |
| HLHS | 0.85 (0.81,0.88) | 0.74 (0.65,0.83) | 0.76 (0.74,0.77) | 3.07 (2.65,3.51) | 0.34 (0.22,0.46) | 5.65 (3.86,7.78) |
| LSVC | 0.71 (0.66,0.75) | 0.65 (0.56,0.75) | 0.63 (0.61,0.65) | 1.78 (1.51,2.04) | 0.55 (0.40,0.70) | 2.18 (1.70,2.85) |
| PAPVC | 0.76 (0.67,0.83) | 0.58 (0.44,0.71) | 0.8 (0.79,0.82) | 2.92 (2.19,3.69) | 0.53 (0.35,0.70) | 8.41 (4.25,14.51) |
| PDA | 0.8 (0.77,0.82) | 0.76 (0.71,0.80) | 0.72 (0.71,0.74) | 2.74 (2.53,3.00) | 0.34 (0.27,0.40) | 3.37 (2.87,3.94) |
| Right Aortic Arch | 0.65 (0.59,0.72) | 0.55 (0.43,0.67) | 0.69 (0.68,0.71) | 1.81 (1.43,2.23) | 0.64 (0.47,0.82) | 2.59 (1.57,4.42) |
| Tricuspid Atresia | 0.76 (0.65,0.87) | 0.28 (0.07,0.50) | 0.95 (0.95,0.96) | 5.91 (1.55,10.91) | 0.76 (0.52,0.97) | 8.78 (2.28,26.03) |
| Truncus arteriosus | 0.77 (0.65,0.87) | 0.57 (0.33,0.79) | 0.78 (0.77,0.80) | 2.65 (1.57,3.67) | 0.55 (0.27,0.85) | 12.19 (2.85,32.18) |
| SV Disease | 0.83 (0.80,0.86) | 0.77 (0.70,0.83) | 0.75 (0.73,0.76) | 3.05 (2.73,3.41) | 0.31 (0.22,0.40) | 4.1 (3.23,5.16) |
| Tetralogy of Fallot | 0.81 (0.77,0.85) | 0.68 (0.58,0.77) | 0.8 (0.78,0.81) | 3.34 (2.82,3.85) | 0.41 (0.29,0.53) | 6.06 (3.91,8.79) |
| AVCD | 0.8 (0.78,0.82) | 0.78 (0.74,0.83) | 0.66 (0.65,0.68) | 2.33 (2.15,2.50) | 0.33 (0.26,0.40) | 3.38 (2.87,3.94) |
| VSD | 0.72 (0.70,0.74) | 0.69 (0.66,0.73) | 0.61 (0.60,0.63) | 1.79 (1.66,1.92) | 0.5 (0.44,0.57) | 2.19 (1.98,2.40) |
| CoA | 0.72 (0.68,0.75) | 0.71 (0.64,0.78) | 0.61 (0.60,0.63) | 1.83 (1.64,2.03) | 0.47 (0.37,0.59) | 2.51 (1.98,3.21) |
| Pulmonary Atresia | 0.72 (0.67,0.76) | 0.61 (0.52,0.71) | 0.7 (0.68,0.71) | 2.04 (1.70,2.35) | 0.55 (0.42,0.69) | 2.29 (1.72,3.16) |
| TAPVC | 0.74 (0.65,0.82) | 0.47 (0.28,0.66) | 0.79 (0.77,0.80) | 2.22 (1.33,3.14) | 0.67 (0.43,0.91) | 7.63 (2.12,19.24) |
| Composite Non-Critical CHD | 0.69 (0.67,0.71) | 0.76 (0.74,0.78) | 0.53 (0.50,0.55) | 1.61 (1.52,1.70) | 0.45 (0.41,0.50) | 1.37 (1.33,1.43) |
| Composite Critical CHD | 0.77 (0.76,0.79) | 0.79 (0.77,0.82) | 0.6 (0.58,0.62) | 2.0 (1.88,2.12) | 0.34 (0.30,0.38) | 1.87 (1.77,1.97) |
| Infant Composite Critical CHD | 0.71 (0.68,0.74) | 0.85 (0.82,0.88) | 0.41 (0.38,0.45) | 1.45 (1.35,1.55) | 0.36 (0.28,0.45) | 1.53 (1.42,1.63) |

### Table S4: External (US) Model Performance for Each Outcome

| | AUROC | Sensitivity | Specificity | LR + | LR - | Lift |
|---|---|---|---|---|---|---|
| ASD | 0.73 (0.71,0.76) | 0.77 (0.73,0.81) | 0.57 (0.55,0.59) | 1.78 (1.65,1.91) | 0.41 (0.34,0.49) | 2.09 (1.88,2.32) |
| Anomalous Coronaries | 0.71 (0.63,0.78) | 0.63 (0.49,0.77) | 0.65 (0.63,0.67) | 1.81 (1.41,2.21) | 0.57 (0.36,0.78) | 4.3 (2.15,7.65) |
| BAV | 0.73 (0.69,0.77) | 0.72 (0.66,0.78) | 0.6 (0.58,0.62) | 1.81 (1.64,2.00) | 0.46 (0.36,0.57) | 3.55 (2.86,4.29) |
| DORV | 0.79 (0.75,0.82) | 0.64 (0.55,0.72) | 0.79 (0.77,0.81) | 3.03 (2.55,3.49) | 0.46 (0.35,0.57) | 3.21 (2.40,4.35) |
| D-loop TGA | 0.68 (0.61,0.75) | 0.67 (0.55,0.80) | 0.57 (0.55,0.59) | 1.58 (1.26,1.87) | 0.57 (0.36,0.80) | 2.99 (1.66,5.33) |
| Ebstein | 0.79 (0.73,0.85) | 0.7 (0.59,0.83) | 0.69 (0.68,0.71) | 2.3 (1.90,2.73) | 0.43 (0.25,0.60) | 7.12 (4.14,11.08) |
| HLHS | 0.81 (0.77,0.85) | 0.73 (0.63,0.83) | 0.72 (0.70,0.74) | 2.64 (2.26,3.02) | 0.37 (0.24,0.51) | 4.3 (3.00,6.15) |
| LSVC | 0.68 (0.62,0.73) | 0.67 (0.57,0.77) | 0.56 (0.54,0.58) | 1.54 (1.29,1.78) | 0.58 (0.40,0.77) | 2.0 (1.53,2.75) |
| PAPVC | 0.75 (0.66,0.84) | 0.63 (0.47,0.78) | 0.76 (0.74,0.77) | 2.58 (1.93,3.21) | 0.49 (0.30,0.70) | 9.08 (4.44,16.24) |
| PDA | 0.78 (0.75,0.82) | 0.76 (0.70,0.81) | 0.69 (0.67,0.71) | 2.46 (2.21,2.71) | 0.35 (0.27,0.44) | 3.44 (2.79,4.19) |
| Right Aortic Arch | 0.65 (0.59,0.71) | 0.58 (0.46,0.69) | 0.66 (0.64,0.68) | 1.7 (1.36,2.04) | 0.64 (0.46,0.81) | 2.14 (1.43,3.45) |
| Tricuspid Atresia | 0.72 (0.56,0.88) | 0.37 (0.11,0.67) | 0.95 (0.94,0.95) | 6.8 (1.94,12.93) | 0.67 (0.35,0.94) | 13.93 (2.50,47.92) |
| Truncus arteriosus | 0.79 (0.69,0.89) | 0.6 (0.36,0.83) | 0.74 (0.72,0.76) | 2.31 (1.37,3.26) | 0.54 (0.22,0.87) | 13.76 (3.36,34.11) |
| SV Disease | 0.81 (0.77,0.84) | 0.77 (0.69,0.85) | 0.71 (0.69,0.73) | 2.69 (2.37,3.01) | 0.32 (0.21,0.43) | 3.66 (2.83,4.79) |
| Tetralogy of Fallot | 0.81 (0.77,0.85) | 0.74 (0.64,0.83) | 0.75 (0.74,0.77) | 3.02 (2.57,3.42) | 0.34 (0.22,0.47) | 5.44 (3.56,7.91) |
| AVCD | 0.78 (0.75,0.81) | 0.81 (0.76,0.86) | 0.6 (0.58,0.63) | 2.06 (1.91,2.23) | 0.31 (0.23,0.39) | 2.89 (2.46,3.39) |
| VSD | 0.73 (0.71,0.76) | 0.72 (0.68,0.76) | 0.59 (0.57,0.62) | 1.79 (1.66,1.93) | 0.46 (0.39,0.54) | 2.1 (1.89,2.32) |
| CoA | 0.7 (0.65,0.74) | 0.72 (0.64,0.79) | 0.56 (0.54,0.58) | 1.62 (1.44,1.81) | 0.51 (0.38,0.64) | 2.53 (1.88,3.49) |
| Pulmonary Atresia | 0.71 (0.66,0.76) | 0.67 (0.57,0.77) | 0.65 (0.63,0.67) | 1.91 (1.61,2.20) | 0.51 (0.36,0.67) | 2.39 (1.67,3.61) |
| TAPVC | 0.74 (0.64,0.83) | 0.5 (0.28,0.72) | 0.75 (0.73,0.77) | 2.01 (1.13,2.88) | 0.66 (0.37,0.96) | 4.71 (1.83,11.32) |
| Composite Non-Critical CHD | 0.65 (0.63,0.67) | 0.78 (0.75,0.80) | 0.44 (0.41,0.47) | 1.39 (1.31,1.47) | 0.51 (0.45,0.57) | 1.25 (1.20,1.29) |
| Composite Critical CHD | 0.74 (0.72,0.76) | 0.81 (0.79,0.84) | 0.52 (0.50,0.55) | 1.71 (1.61,1.83) | 0.36 (0.30,0.41) | 1.66 (1.56,1.76) |
| Infant Composite Critical CHD | 0.68 (0.64,0.71) | 0.87 (0.84,0.90) | 0.32 (0.28,0.36) | 1.28 (1.20,1.37) | 0.41 (0.30,0.53) | 1.38 (1.28,1.49) |

**Table S5: External (International) Model Performance for Each Outcome**

| | AUROC | Sensitivity | Specificity | LR + | LR - | Lift |
|---|---|---|---|---|---|---|
| ASD | 0.74 (0.69,0.79) | 0.6 (0.50,0.70) | 0.73 (0.70,0.75) | 2.2 (1.78,2.60) | 0.55 (0.42,0.69) | 2.52 (1.93,3.24) |
| Anomalous Coronaries | 0.79 (0.52,0.96) | 0.67 (0.20,1.00) | 0.8 (0.77,0.83) | 3.36 (1.05,5.27) | 0.41 (0.00,0.99) | 27.4 (2.13,117.26) |
| BAV | 0.8 (0.73,0.86) | 0.63 (0.46,0.78) | 0.75 (0.72,0.78) | 2.52 (1.82,3.26) | 0.5 (0.29,0.72) | 7.21 (3.66,12.02) |
| DORV | 0.83 (0.78,0.88) | 0.3 (0.14,0.48) | 0.9 (0.88,0.92) | 3.0 (1.40,4.93) | 0.78 (0.58,0.95) | 4.19 (2.52,7.34) |
| D-loop TGA | 0.76 (0.66,0.85) | 0.53 (0.30,0.76) | 0.77 (0.74,0.79) | 2.3 (1.26,3.32) | 0.61 (0.30,0.91) | 3.66 (1.85,7.99) |
| Ebstein | 0.78 (0.64,0.90) | 0.64 (0.40,0.87) | 0.79 (0.77,0.82) | 3.1 (1.90,4.23) | 0.45 (0.16,0.76) | 8.47 (2.66,19.80) |
| HLHS | 0.93 (0.90,0.96) | 0.79 (0.62,0.95) | 0.84 (0.82,0.86) | 5.06 (3.91,6.32) | 0.24 (0.06,0.45) | 13.73 (7.32,23.95) |
| LSVC | 0.74 (0.59,0.87) | 0.53 (0.25,0.80) | 0.79 (0.76,0.81) | 2.54 (1.18,3.92) | 0.59 (0.25,0.95) | 3.34 (1.71,6.51) |
| PAPVC | 0.8 (0.65,0.93) | 0.4 (0.10,0.73) | 0.91 (0.89,0.93) | 4.53 (1.06,8.41) | 0.66 (0.30,0.99) | 5.24 (2.25,10.21) |
| PDA | 0.83 (0.78,0.87) | 0.75 (0.67,0.83) | 0.8 (0.77,0.83) | 3.8 (3.15,4.52) | 0.31 (0.20,0.42) | 3.54 (2.79,4.46) |
| Right Aortic Arch | 0.48 (0.23,0.72) | 0.33 (0.00,0.67) | 0.78 (0.75,0.81) | 1.5 (0.00,2.95) | 0.86 (0.43,1.29) | 11.36 (0.80,42.49) |
| Tricuspid Atresia | 0.86 (0.74,0.94) | 0.13 (0.00,0.50) | 0.97 (0.96,0.98) | 4.67 (0.00,16.76) | 0.89 (0.52,1.04) | 6.89 (2.94,16.91) |
| Truncus arteriosus | 0.66 (0.11,0.95) | 0.51 (0.00,1.00) | 0.88 (0.86,0.90) | 4.4 (0.00,9.16) | 0.56 (0.00,1.14) | 4.93 (1.12,19.43) |
| SV Disease | 0.89 (0.85,0.93) | 0.75 (0.62,0.88) | 0.83 (0.81,0.86) | 4.56 (3.59,5.64) | 0.29 (0.15,0.45) | 7.01 (4.51,10.63) |
| Tetralogy of Fallot | 0.75 (0.64,0.85) | 0.31 (0.08,0.53) | 0.9 (0.88,0.92) | 3.07 (0.80,5.49) | 0.77 (0.53,1.02) | 8.97 (1.76,22.93) |
| AVCD | 0.78 (0.70,0.86) | 0.57 (0.41,0.74) | 0.79 (0.76,0.81) | 2.73 (1.95,3.61) | 0.54 (0.33,0.74) | 6.12 (3.16,10.44) |
| VSD | 0.64 (0.58,0.69) | 0.55 (0.45,0.65) | 0.65 (0.62,0.68) | 1.56 (1.24,1.88) | 0.7 (0.54,0.86) | 1.94 (1.39,2.63) |
| CoA | 0.75 (0.67,0.84) | 0.66 (0.45,0.88) | 0.74 (0.71,0.76) | 2.52 (1.73,3.46) | 0.46 (0.17,0.74) | 3.14 (1.77,5.77) |
| Pulmonary Atresia | 0.74 (0.66,0.81) | 0.42 (0.23,0.61) | 0.82 (0.80,0.84) | 2.34 (1.24,3.43) | 0.71 (0.48,0.95) | 2.3 (1.59,3.42) |
| TAPVC | 0.73 (0.50,0.91) | 0.38 (0.00,0.75) | 0.88 (0.86,0.90) | 3.21 (0.00,6.41) | 0.71 (0.28,1.14) | 17.72 (1.69,67.34) |
| Composite Non-Critical CHD | 0.74 (0.71,0.77) | 0.7 (0.65,0.75) | 0.67 (0.63,0.70) | 2.11 (1.85,2.39) | 0.45 (0.37,0.53) | 1.79 (1.61,1.98) |
| Composite Critical CHD | 0.82 (0.79,0.85) | 0.71 (0.65,0.78) | 0.75 (0.72,0.78) | 2.86 (2.43,3.33) | 0.39 (0.29,0.47) | 2.61 (2.26,3.05) |
| Infant Composite Critical CHD | 0.73 (0.67,0.79) | 0.74 (0.64,0.84) | 0.6 (0.54,0.65) | 1.85 (1.51,2.23) | 0.43 (0.27,0.61) | 2.13 (1.69,2.62) |

### Table S6: External Model Performance Using LLM-Derived Labels

| | AUROC | Sensitivity | Specificity | LR + | LR - | Lift |
|---|---|---|---|---|---|---|
| ASD | 0.79 (0.77,0.81) | 0.76 (0.72,0.81) | 0.67 (0.65,0.69) | 2.32 (2.13,2.51) | 0.35 (0.29,0.42) | 2.65 (2.38,2.97) |
| Anomalous Coronaries | 0.75 (0.67,0.82) | 0.63 (0.49,0.77) | 0.68 (0.66,0.70) | 1.99 (1.51,2.43) | 0.54 (0.33,0.76) | 6.76 (2.82,12.30) |
| BAV | 0.76 (0.71,0.80) | 0.67 (0.60,0.75) | 0.69 (0.67,0.71) | 2.18 (1.93,2.46) | 0.47 (0.36,0.58) | 4.19 (3.19,5.34) |
| DORV | 0.84 (0.79,0.89) | 0.52 (0.33,0.69) | 0.9 (0.88,0.91) | 5.03 (3.34,6.74) | 0.54 (0.35,0.74) | 7.31 (3.13,15.25) |
| D-loop TGA | 0.72 (0.62,0.80) | 0.63 (0.43,0.82) | 0.73 (0.71,0.75) | 2.33 (1.58,3.04) | 0.5 (0.25,0.78) | 5.35 (1.55,15.32) |
| Ebstein | 0.84 (0.77,0.89) | 0.76 (0.65,0.87) | 0.75 (0.73,0.76) | 3.01 (2.51,3.50) | 0.32 (0.17,0.48) | 8.81 (5.10,13.16) |
| HLHS | 0.87 (0.78,0.94) | 0.67 (0.38,0.92) | 0.86 (0.84,0.87) | 4.72 (2.61,6.69) | 0.38 (0.10,0.72) | 27.54 (3.81,81.18) |
| LSVC | 0.73 (0.65,0.80) | 0.63 (0.48,0.76) | 0.69 (0.67,0.71) | 2.02 (1.53,2.46) | 0.54 (0.34,0.76) | 3.29 (1.99,5.69) |
| PAPVC | 0.82 (0.75,0.89) | 0.68 (0.52,0.81) | 0.8 (0.78,0.81) | 3.35 (2.57,4.12) | 0.41 (0.23,0.60) | 9.55 (4.99,15.68) |
| PDA | 0.8 (0.77,0.83) | 0.76 (0.71,0.81) | 0.72 (0.70,0.74) | 2.74 (2.48,3.01) | 0.33 (0.26,0.40) | 3.05 (2.63,3.58) |
| Right Aortic Arch | 0.68 (0.59,0.76) | 0.5 (0.36,0.65) | 0.76 (0.75,0.78) | 2.12 (1.51,2.77) | 0.65 (0.46,0.84) | 4.44 (1.94,8.92) |
| Tricuspid Atresia | 0.79 (0.56,0.97) | 0.17 (0.00,0.50) | 0.98 (0.97,0.99) | 8.62 (0.00,30.19) | 0.85 (0.51,1.02) | 48.66 (2.05,265.03) |
| Truncus arteriosus | 0.55 (0.06,1.00) | 0.34 (0.00,1.00) | 0.86 (0.85,0.88) | 2.51 (0.00,7.60) | 0.76 (0.00,1.17) | 104.07 (0.98,589.50) |
| SV Disease | 0.86 (0.80,0.92) | 0.74 (0.58,0.89) | 0.85 (0.83,0.86) | 4.87 (3.76,5.98) | 0.31 (0.13,0.50) | 8.1 (4.14,15.38) |
| Tetralogy of Fallot | 0.84 (0.79,0.88) | 0.65 (0.52,0.76) | 0.85 (0.84,0.86) | 4.33 (3.43,5.26) | 0.41 (0.28,0.56) | 8.25 (4.83,12.66) |
| AVCD | 0.84 (0.81,0.87) | 0.79 (0.72,0.85) | 0.74 (0.72,0.75) | 3.0 (2.69,3.32) | 0.29 (0.20,0.38) | 4.31 (3.45,5.51) |
| VSD | 0.76 (0.73,0.79) | 0.74 (0.69,0.78) | 0.63 (0.61,0.65) | 2.0 (1.83,2.18) | 0.42 (0.35,0.49) | 2.63 (2.33,2.98) |
| CoA | 0.78 (0.73,0.82) | 0.73 (0.64,0.82) | 0.69 (0.67,0.71) | 2.37 (2.05,2.70) | 0.39 (0.26,0.53) | 3.7 (2.63,5.16) |
| Pulmonary Atresia | 0.78 (0.71,0.85) | 0.55 (0.35,0.74) | 0.78 (0.77,0.80) | 2.56 (1.58,3.51) | 0.57 (0.33,0.84) | 6.02 (2.37,14.78) |
| TAPVC | 0.88 (0.81,0.94) | 0.58 (0.29,0.88) | 0.84 (0.83,0.86) | 3.68 (1.75,5.53) | 0.5 (0.15,0.86) | 26.88 (4.03,88.27) |
| Composite Non-Critical CHD | 0.72 (0.70,0.74) | 0.77 (0.75,0.80) | 0.58 (0.55,0.61) | 1.84 (1.72,1.98) | 0.39 (0.35,0.44) | 1.44 (1.38,1.50) |
| Composite Critical CHD | 0.79 (0.77,0.80) | 0.69 (0.66,0.73) | 0.72 (0.69,0.74) | 2.43 (2.23,2.64) | 0.43 (0.38,0.48) | 1.99 (1.86,2.13) |
| Infant Composite Critical CHD | 0.72 (0.69,0.75) | 0.81 (0.77,0.84) | 0.49 (0.44,0.53) | 1.59 (1.45,1.75) | 0.39 (0.31,0.47) | 1.47 (1.38,1.57) |

**Table S7: Model Performance by Number of Videos in a Study**

| | Non-Critical CHD AUROC | | Critical CHD AUROC | |
|---|---|---|---|---|
| # of videos | Internal | External | Internal | External |
| <25 | 0.83 (0.77,0.87) | 0.62 (0.58,0.68) | 0.82 (0.70,0.92) | 0.67 (0.62,0.71) |
| 26-50 | 0.84 (0.81,0.88) | 0.65 (0.62,0.69) | 0.94 (0.91,0.96) | 0.74 (0.71,0.77) |
| 51-75 | 0.89 (0.87,0.91) | 0.61 (0.57,0.65) | 0.94 (0.92,0.96) | 0.75 (0.72,0.78) |
| 76-100 | 0.88 (0.87,0.89) | 0.65 (0.58,0.71) | 0.93 (0.90,0.95) | 0.76 (0.70,0.81) |
| >100 | 0.89 (0.87,0.90) | 0.61 (0.52,0.68) | 0.9 (0.89,0.92) | 0.73 (0.65,0.79) |

**Table S8: Model Performance by Age Subgroup**

| | Non-Critical CHD AUROC | | Critical CHD AUROC | |
|---|---|---|---|---|
| Age (years) | Internal | External | Internal | External |
| All ages | 0.9 (0.89,0.91) | 0.69 (0.67,0.71) | 0.94 (0.94,0.95) | 0.77 (0.76,0.79) |
| <1 | 0.86 (0.85,0.87) | 0.61 (0.58,0.64) | 0.93 (0.91,0.94) | 0.71 (0.68,0.73) |
| 1-3 | 0.84 (0.80,0.88) | 0.63 (0.58,0.67) | 0.93 (0.86,0.98) | 0.72 (0.67,0.76) |
| 3-8 | 0.84 (0.81,0.87) | 0.65 (0.61,0.70) | 0.94 (0.91,0.97) | 0.79 (0.75,0.82) |
| 8-12 | 0.78 (0.72,0.83) | 0.71 (0.64,0.77) | 0.96 (0.91,0.99) | 0.83 (0.78,0.88) |
| 12-18 | 0.73 (0.69,0.78) | 0.73 (0.68,0.79) | 0.89 (0.82,0.95) | 0.87 (0.83,0.90) |
| >18 | 0.81 (0.77,0.85) | 0.65 (0.55,0.75) | 0.93 (0.91,0.95) | 0.65 (0.54,0.76) |

**Table S9: Internal Model Performance After Re-Training**

| | AUROC | Sensitivity | Specificity | LR + | LR - | Lift |
|---|---|---|---|---|---|---|
| ASD | 0.87 (0.86,0.88) | 0.81 (0.78,0.83) | 0.79 (0.78,0.80) | 3.91 (3.71,4.12) | 0.24 (0.21,0.28) | 5.61 (5.12,6.18) |
| Anomalous Coronaries | 0.76 (0.70,0.82) | 0.75 (0.65,0.86) | 0.68 (0.67,0.69) | 2.37 (2.04,2.70) | 0.36 (0.21,0.52) | 4.42 (2.65,7.46) |
| BAV | 0.85 (0.82,0.87) | 0.74 (0.69,0.79) | 0.81 (0.80,0.82) | 3.96 (3.62,4.28) | 0.32 (0.25,0.39) | 12.82 (10.63,15.22) |
| DORV | 0.98 (0.97,0.99) | 0.91 (0.81,1.00) | 0.94 (0.93,0.94) | 14.6 (12.69,16.35) | 0.09 (0.00,0.20) | 56.75 (31.49,95.15) |
| D-loop TGA | 0.98 (0.97,0.98) | 0.98 (0.94,1.00) | 0.88 (0.87,0.88) | 7.92 (7.43,8.36) | 0.02 (0.00,0.06) | 49.47 (32.85,71.65) |
| Ebstein | 0.98 (0.96,0.99) | 0.88 (0.72,1.00) | 0.93 (0.93,0.93) | 12.62 (10.34,14.47) | 0.13 (0.00,0.30) | 142.91 (68.12,249.01) |
| HLHS | 0.99 (0.99,1.00) | 0.95 (0.88,1.00) | 0.97 (0.96,0.97) | 28.08 (24.67,31.67) | 0.05 (0.00,0.12) | 132.48 (91.30,189.59) |
| LSVC | 0.9 (0.87,0.92) | 0.82 (0.76,0.88) | 0.8 (0.79,0.81) | 4.13 (3.78,4.47) | 0.22 (0.14,0.30) | 13.26 (9.54,18.00) |
| PAPVC | 0.85 (0.80,0.90) | 0.63 (0.51,0.74) | 0.88 (0.88,0.89) | 5.32 (4.25,6.33) | 0.42 (0.29,0.56) | 19.75 (10.80,32.13) |
| PDA | 0.96 (0.95,0.97) | 0.9 (0.89,0.92) | 0.9 (0.89,0.90) | 8.64 (8.13,9.19) | 0.11 (0.09,0.13) | 7.07 (6.74,7.45) |
| Right Aortic Arch | 0.86 (0.82,0.90) | 0.77 (0.69,0.85) | 0.83 (0.83,0.84) | 4.63 (4.11,5.15) | 0.28 (0.18,0.38) | 16.77 (10.33,24.44) |
| Tricuspid Atresia | 1.0 (0.99,1.00) | 1.0 (1.00,1.00) | 0.97 (0.97,0.97) | 32.4 (29.13,35.97) | 0.0 (0.00,0.00) | 534.53 (284.78,995.45) |
| Truncus arteriosus | 0.96 (0.93,0.98) | 0.92 (0.71,1.00) | 0.86 (0.85,0.87) | 6.54 (5.12,7.36) | 0.09 (0.00,0.33) | 96.18 (16.10,275.78) |
| SV Disease | 0.99 (0.99,1.00) | 1.0 (1.00,1.00) | 0.94 (0.93,0.94) | 16.47 (15.36,17.68) | 0.0 (0.00,0.00) | 91.84 (69.46,118.35) |
| Tetralogy of Fallot | 0.98 (0.97,0.99) | 0.95 (0.91,0.99) | 0.92 (0.91,0.92) | 11.81 (10.90,12.71) | 0.05 (0.01,0.10) | 51.82 (41.32,64.83) |
| AVCD | 0.97 (0.95,0.98) | 0.94 (0.90,0.98) | 0.87 (0.87,0.88) | 7.4 (6.90,7.85) | 0.06 (0.02,0.12) | 44.78 (35.04,57.50) |
| VSD | 0.91 (0.90,0.92) | 0.87 (0.84,0.89) | 0.78 (0.77,0.79) | 4.02 (3.84,4.21) | 0.17 (0.14,0.20) | 7.37 (6.84,7.97) |
| CoA | 0.89 (0.87,0.91) | 0.83 (0.77,0.87) | 0.77 (0.76,0.77) | 3.52 (3.27,3.76) | 0.23 (0.16,0.30) | 15.96 (12.65,19.61) |
| Pulmonary Atresia | 0.97 (0.95,0.98) | 0.87 (0.78,0.96) | 0.9 (0.89,0.90) | 8.44 (7.43,9.46) | 0.14 (0.05,0.24) | 57.25 (35.84,83.67) |
| TAPVC | 0.95 (0.91,0.99) | 0.7 (0.40,0.92) | 0.95 (0.94,0.95) | 13.31 (7.81,18.48) | 0.32 (0.08,0.63) | 131.93 (17.60,373.55) |
| Composite Non-Critical CHD | 0.9 (0.89,0.91) | 0.78 (0.76,0.80) | 0.86 (0.86,0.87) | 5.69 (5.37,6.04) | 0.25 (0.24,0.27) | 3.35 (3.25,3.46) |
| Composite Critical CHD | 0.95 (0.94,0.96) | 0.88 (0.86,0.90) | 0.89 (0.88,0.89) | 7.84 (7.34,8.33) | 0.13 (0.11,0.16) | 11.02 (10.16,11.91) |
| Infant Composite Critical CHD | 0.93 (0.91,0.94) | 0.92 (0.90,0.95) | 0.75 (0.73,0.76) | 3.67 (3.42,3.94) | 0.1 (0.07,0.14) | 6.08 (5.56,6.67) |

**Table S10: External (International) Model Performance After Re-Training**

| | AUROC | Sensitivity | Specificity | LR + | LR - | Lift |
|---|---|---|---|---|---|---|
| ASD | 0.74 (0.69,0.79) | 0.7 (0.61,0.78) | 0.68 (0.64,0.71) | 2.17 (1.83,2.55) | 0.44 (0.32,0.58) | 2.25 (1.85,2.75) |
| Anomalous Coronaries | 0.63 (0.48,0.78) | 0.72 (0.50,0.94) | 0.4 (0.36,0.44) | 1.2 (0.82,1.58) | 0.7 (0.15,1.28) | 7.04 (1.35,17.90) |
| BAV | 0.64 (0.54,0.75) | 0.57 (0.41,0.73) | 0.63 (0.60,0.66) | 1.56 (1.11,2.03) | 0.68 (0.43,0.94) | 4.69 (2.25,7.87) |
| DORV | 0.93 (0.88,0.96) | 0.9 (0.67,1.00) | 0.85 (0.82,0.87) | 5.92 (4.27,7.42) | 0.12 (0.00,0.39) | 13.53 (5.30,33.20) |
| D-loop TGA | 0.8 (0.68,0.94) | 0.41 (0.00,1.00) | 0.74 (0.71,0.77) | 1.58 (0.00,3.82) | 0.8 (0.00,1.38) | 5.07 (2.27,13.79) |
| Ebstein | 0.95 (0.87,1.00) | 0.91 (0.71,1.00) | 0.77 (0.74,0.80) | 3.98 (2.97,4.82) | 0.12 (0.00,0.38) | 40.41 (19.22,85.45) |
| HLHS | 0.92 (0.81,0.98) | 0.67 (0.33,1.00) | 0.94 (0.92,0.95) | 10.93 (5.44,17.36) | 0.35 (0.00,0.71) | 19.85 (6.83,48.51) |
| LSVC | 0.62 (0.51,0.73) | 0.49 (0.31,0.67) | 0.72 (0.69,0.75) | 1.73 (1.09,2.40) | 0.72 (0.46,0.96) | 2.04 (1.24,3.48) |
| PAPVC | 0.86 (0.76,0.94) | 0.63 (0.31,0.92) | 0.89 (0.87,0.91) | 5.77 (2.77,8.87) | 0.41 (0.09,0.77) | 6.97 (3.18,13.40) |
| PDA | 0.84 (0.78,0.88) | 0.74 (0.65,0.82) | 0.8 (0.76,0.83) | 3.66 (2.99,4.44) | 0.32 (0.22,0.44) | 3.85 (3.14,4.80) |
| Right Aortic Arch | 0.69 (0.47,0.89) | 0.5 (0.14,0.88) | 0.71 (0.68,0.74) | 1.75 (0.49,3.18) | 0.7 (0.17,1.23) | 8.78 (1.41,31.75) |
| Tricuspid Atresia | 0.93 (0.86,0.98) | 0.41 (0.00,1.00) | 0.97 (0.96,0.98) | 13.52 (0.00,32.89) | 0.61 (0.00,1.04) | 18.73 (5.93,51.50) |
| Truncus arteriosus | 0.48 (0.14,0.83) | 0.49 (0.00,1.00) | 0.76 (0.73,0.79) | 2.08 (0.00,4.59) | 0.66 (0.00,1.34) | 2.62 (1.16,5.89) |
| SV Disease | 0.92 (0.88,0.96) | 0.56 (0.33,0.79) | 0.89 (0.87,0.91) | 5.17 (2.93,7.53) | 0.5 (0.24,0.75) | 17.42 (8.16,32.64) |
| Tetralogy of Fallot | 0.8 (0.72,0.88) | 0.32 (0.07,0.62) | 0.84 (0.82,0.87) | 2.06 (0.42,4.04) | 0.8 (0.45,1.12) | 3.47 (2.08,6.27) |
| AVCD | 0.87 (0.79,0.94) | 0.83 (0.67,0.96) | 0.77 (0.74,0.80) | 3.58 (2.76,4.42) | 0.22 (0.05,0.44) | 10.49 (5.43,17.96) |
| VSD | 0.72 (0.67,0.78) | 0.84 (0.76,0.91) | 0.45 (0.41,0.48) | 1.51 (1.35,1.69) | 0.37 (0.21,0.54) | 2.6 (2.01,3.40) |
| CoA | 0.8 (0.71,0.89) | 0.84 (0.64,1.00) | 0.61 (0.58,0.65) | 2.17 (1.63,2.63) | 0.27 (0.00,0.59) | 7.46 (2.63,15.98) |
| Pulmonary Atresia | 0.88 (0.85,0.92) | 1.0 (1.00,1.00) | 0.82 (0.79,0.85) | 5.57 (4.79,6.49) | 0.0 (0.00,0.00) | 5.29 (3.99,8.11) |
| TAPVC | 0.88 (0.74,0.99) | 0.71 (0.33,1.00) | 0.92 (0.90,0.94) | 9.22 (3.70,14.25) | 0.31 (0.00,0.74) | 35.11 (4.32,102.31) |
| Composite Non-Critical CHD | 0.72 (0.69,0.76) | 0.77 (0.73,0.82) | 0.52 (0.47,0.57) | 1.61 (1.44,1.80) | 0.44 (0.34,0.54) | 1.65 (1.52,1.80) |
| Composite Critical CHD | 0.87 (0.83,0.90) | 0.86 (0.80,0.92) | 0.72 (0.69,0.76) | 3.15 (2.70,3.63) | 0.19 (0.10,0.28) | 3.92 (3.20,4.74) |
| Infant Composite Critical CHD | 0.84 (0.79,0.89) | 0.96 (0.91,1.00) | 0.48 (0.41,0.54) | 1.83 (1.60,2.10) | 0.09 (0.00,0.20) | 2.54 (2.09,3.13) |

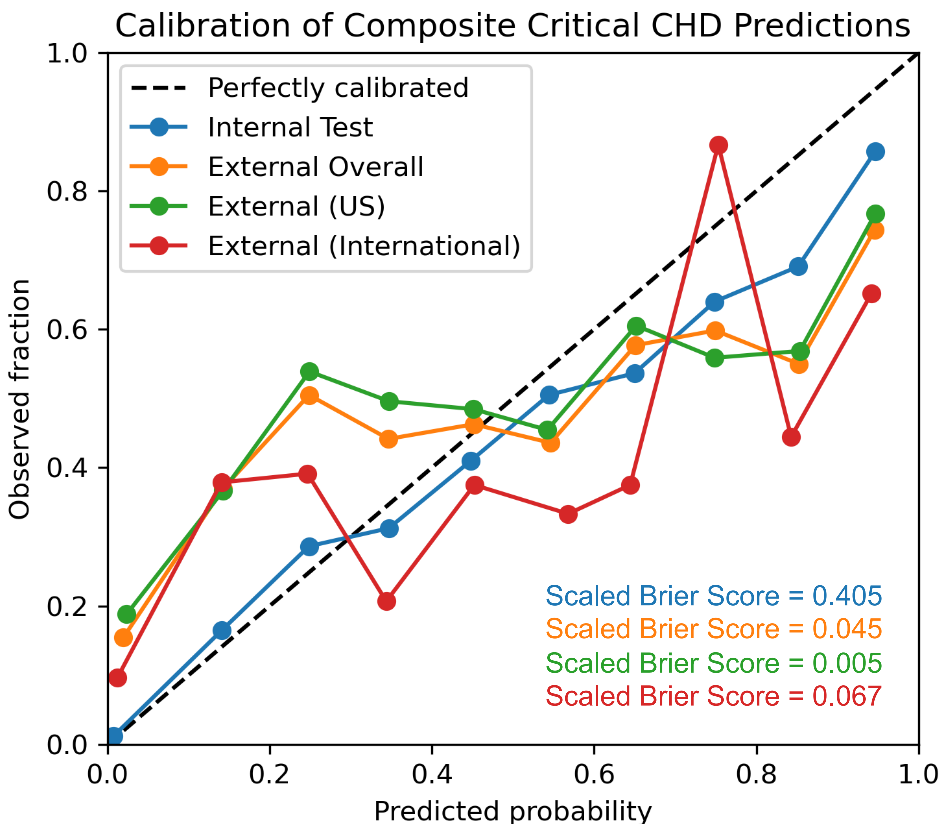

**Figure S1: Calibration Plots.** Calibration plots of the internal (blue), overall external (orange), external US (green), and external international (red) cohorts. Scaled brier scores inset.

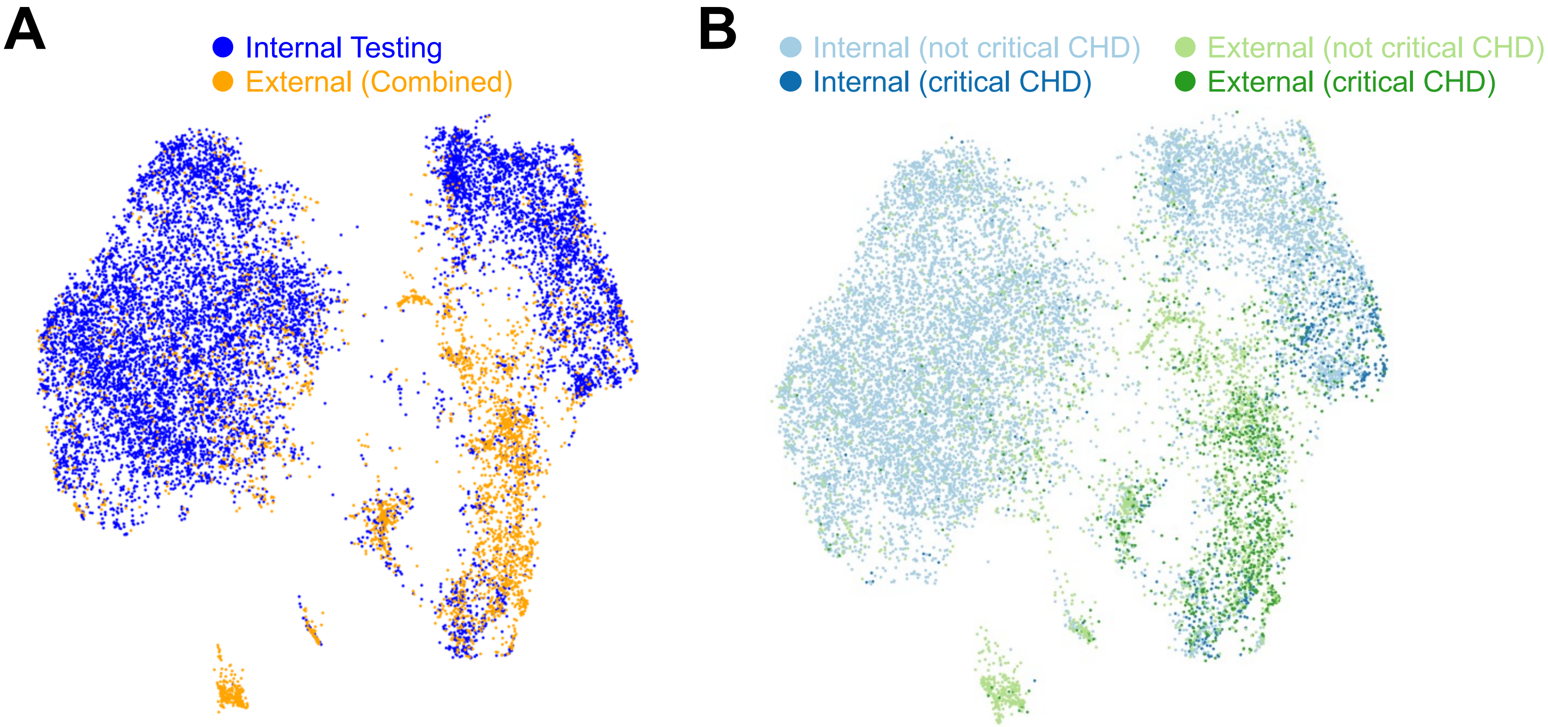


**Figure S2: UMAP Visualization of Study-Level Embeddings.** (**A**) Visualization of internal (blue) and external (orange) echo study embeddings projected onto two dimensions using UMAP. (**B**) Stratifying internal and external embeddings by the presence (dark) or absence (light) of critical CHD.

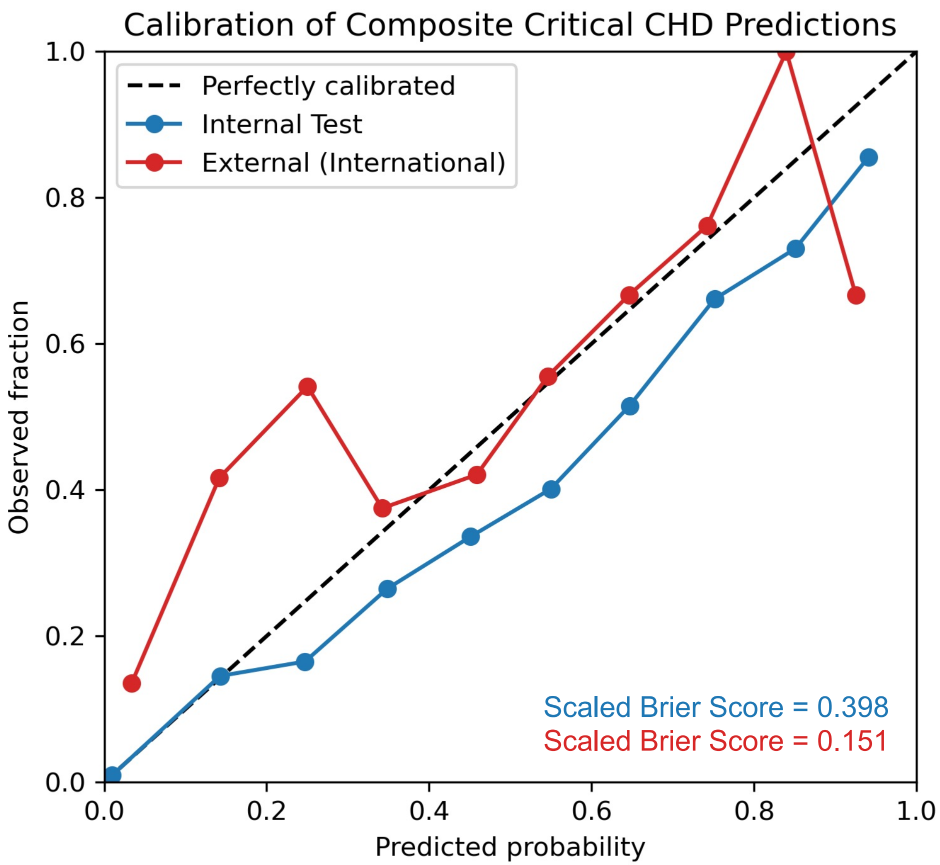


**Figure S3: Calibration Plots After Re-Training.** Calibration plots of the internal (blue) and external international (red) cohorts. Scaled brier scores inset.
